# Meta-analysis of genome-wide association studies of doxorubicin-induced arrhythmia identifies *MMP16 and KCTD15 as risk loci*

**DOI:** 10.64898/2025.12.16.25342433

**Authors:** Joseph S. Reddy, Milagros Pereyra, Mohammed Tiseer Abbas, Reza Arsanjani, Isaac E. Prah, Nicholas J. Boddicker, James R. Cerhan, Melissa C. Larson, Raphael Mwangi, Thomas M Habermann, Hector R. Villarraga, Robert A. Vierkant, Tony C. Luehrs, Hugues Sicotte, Jan B. Egan, Konstantinos N. Lazaridis, Maryam Moossavi, Xiaolei Xu, Pooja P. Advani, Chadi Ayoub, Nadine Norton

**Affiliations:** Quantitative Health Sciences, Mayo Clinic Florida, Jacksonville, FL, USA; Cardiovascular Medicine, Mayo Clinic Arizona, Scottsdale, AZ, USA; Cancer Biology, Mayo Clinic Florida, Jacksonville, FL, USA; Division of Computational Biology, Mayo Clinic, Rochester, MN, USA; Division of Epidemiology, Mayo Clinic, Rochester, MN, USA; Division of Clinical Trials and Biostatistics, Mayo Clinic, Rochester, MN, USA; Division of Hematology, Mayo Clinic, Rochester, MN, USA; Department of Cardiovascular Medicine, Mayo Clinic College of Medicine and Science, Rochester, MN, USA; Quantitative Health Sciences, Mayo Clinic, Rochester, MN, USA; Center for Individualized Medicine, Mayo Clinic, Scottsdale, AZ, USA; Division of Gastroenterology and Hepatology, College of Medicine, Mayo Clinic, Rochester, MN, USA; Biochemistry and Molecular Biology, Mayo Clinic, Rochester, MN, USA; Hematology and Oncology, Mayo Clinic Florida, Jacksonville, FL, USA

**Keywords:** Anthracycline, adriamycin, cardiomyopathy, cardiotoxicity, heart failure, chemotherapy-related heart failure, metalloproteinase, potassium channel

## Abstract

**Background:** Anthracyclines are a widely used and effective class of chemotherapy. However, a major limitation for their use is cardiotoxicity, manifesting as systolic dysfunction, congestive heart failure (HF) and arrhythmias.

**Objective:** Identify genes and genetic risk variants for doxorubicin-induced arrhythmia.

**Methods:** We performed genome-wide association studies (GWAS) and meta-analysis across two independent non-overlapping, genetically homogenous datasets from the Molecular Epidemiology Resource (MER, cases N=77, controls, N=1,184) and Tapestry, (cases N=13, controls, N=172). Doxorubicin-related arrhythmia was the primary cardiac outcome followed by investigation of a broader phenotype of cardiac events. Effects of doxorubicin on top associated genes were assessed by qPCR on RNA extracted from human cardiomyocytes following treatment with doxorubicin and by echocardiography in a zebrafish model of doxorubicin-induced cardiomyopathy.

**Results:** In the meta-analysis, *KCTD15* rs12151014 was associated with doxorubicin-related arrhythmia at the genome-wide significance, (MAF in MER cases 0.195 vs 0.069 in controls; Tapestry cases 0.192 vs 0.079 in controls) p=9.01×10^-9^, OR 3.61. *MMP*16 rs1094199, was associated with the broader phenotype of arrhythmia and/or HF (MAF in MER cases 0.08 vs 0.01 in controls; Tapestry cases 0.038 vs 0.015) p = 1.98 x 10^-8^, OR = 6.15. In human cardiomyocytes, *MMP16* and *KCTD15* gene expression was significantly down-regulated by doxorubicin and in a zebrafish *MMP16* knockout was protective of doxorubicin-induced decline in LVEF.

**Conclusions:** Genetic variants at *KCTD15* and *MMP16* are associated with doxorubicin-related arrhythmia and cardiac events. *KCTD15* and *MMP16* are potential cardioprotective therapeutic targets.

## INTRODUCTION

Anthracyclines are a widely used class of chemotherapy that are associated with cytotoxicity through double-strand DNA breaks. This class of chemotherapy is highly effective and is used for treatment of several malignancies including lymphoma, leukemia, Wilms’ tumor, and breast, small cell lung, endometrial, gastric, liver, kidney, ovarian, thyroid, and pediatric cancers.^1–23^ Exposure to anthracyclines is most common in patients with breast cancer and lymphoma. The major limitation for use is cardiotoxicity,^24–27^ manifesting as systolic dysfunction, congestive heart failure (CHF) and arrhythmias including atrial fibrillation, ventricular tachycardia and heart block.^28–30^ The incidence and timing of arrhythmias in patients with or without cardiomyopathy are not well known. ^30^ Although anthracyclines are known to be pro-arrhythmogenic,^29,30^ safety analyses in clinical trials have been primarily focused on outcomes of CHF and left ventricular systolic dysfunction specifically, often overlooking arrhythmia-specific endpoints.^31–35^ Nonetheless, reporting of arrhythmia is important, as atrial fibrillation is associated with stroke risk and generally requires anticoagulation, and ventricular arrhythmias and advanced heart block may be life-threatening.

The Molecular Epidemiology Resource (MER) prospective observational cohort study of newly diagnosed lymphoma characterized the cumulative incidence and risk factors of cardiovascular disease (CVD) in lymphoma patients.^24^ The MER study reported increasing age, male sex, current smoking, body mass index (BMI) and any anthracycline treatment to be significantly associated with risk of cardiovascular disease (CVD) and anthracyclines were associated with increased risk of CHF (HR = 2.71, p < 0.001) and arrhythmia (HR = 1.61, p < 0.01), but not with valvular heart disease or acute coronary syndrome after adjustment for CVD risk factors. Inter-individual variability in anthracycline-related cardiotoxicities is not fully explained by age and other baseline risk factors, suggesting a genetic risk component.^36^

A number of other, non-anthracycline related cardiomyopathies, channelopathies and arrhythmias are known to have a genetic component^37–39^, and the architecture is complex^40–42^. As such, we hypothesized that genome-wide association analysis (GWAS) could identify novel risk genes and variants for anthracycline-induced arrhythmia. Defining such genes has clinical importance, allowing for a personalized medicine approach to identify high-risk individuals for cardiac complications, who may then be better targeted for preventative measures, cardiac optimization, and close surveillance.

Barriers to GWAS of anthracycline-induced arrhythmia are the lack of cohorts with clinical data on this outcome and with corresponding genomic data. However, the MER study,^24^ after excluding prevalent CVD at lymphoma diagnosis, consisted of 3,063 patients, 164 arrhythmia events and availability of SNP data for GWAS analysis. Similarly, the Tapestry study enrolled >98,000 participants at Mayo Clinic, consented for genetic studies as a resource for research discovery. ^43,44^ Therefore, in the present study, we accessed both cohorts to perform GWAS and meta-analysis, specifically for anthracycline-related cardiac events including arrhythmia.

## METHODS

### Patient populations

Clinical and genetic data were obtained through the Molecular Epidemiology Resource (MER)^24^ and Tapestry^43,45^ studies.

The MER study characterized the cumulative incidence and risk factors for cardiovascular disease, consisting of congestive heart failure, acute coronary syndrome, valvular heart disease and arrhythmia, in 3,063 newly diagnosed lymphoma patients (>18 years) who were enrolled at the Mayo Clinic between 2002 and 2015. Genetic data was available for 2,592 MER participants (Supplementary Figure 1). Genotyping of MER participants was obtained through Project Generation, a collaboration between the Mayo Clinic (Center for Individualized Medicine and Mayo Investigators) and Regeneron Genetics Center (RGC) to sequence over 100,000 Mayo Clinic participants from the Mayo Clinic Biobank and research registries, all under a research IRB. Genetic data including whole exome sequencing (WES), Genotyping by Sequencing (GxS) SNP data, as well as imputation was provided by Regeneron. The Regeneron sequencing assay is a heavily modified version of the TWIST Comprehensive Exome custom capture. The custom design included additional augmentation of the exome capture with “backbone” regions intended to capture common tagging variation for purposes of genome wide association studies (GWAS).

Tapestry^43,45^ is a large-scale, decentralized and clinical grade genomic sequencing study that recruited from patients receiving care at Mayo Clinic. Tapestry enrolled >98,000 participants between July 2020 and May 2024. Exome+® sequencing^46,47^ was conducted at Helix, Inc. (San Mateo, CA) with CDC Tier 1 genomic results returned to participants and raw data stored at Mayo Clinic for future research use. The goal of Tapestry was to advance clinical practice, research discovery and genomic education. Written informed consent was obtained for all participants and approved by the Mayo Clinic IRB. In the current study, Tapestry patients with diagnoses of lymphoma or breast cancer and available Exome+® data who received anthracycline therapy were identified (Supplementary Figure 1). These patients were then curated by a team of cardiologists and internal medicine physicians (MP, MA, RA, CA) for demographic data (age at anthracycline dosing, sex, Body Mass Index and diabetes) and cardiac events of heart failure or arrhythmia (atrial fibrillation [AF], ventricular tachycardia [VT and complete heart block [CHB]) that were attributed to anthracycline treatment. AF was defined based on presence of irregular R-R intervals with absence of distinct P waves and presence of fibrillatory waves on standard 12-lead electrocardiogram (ECG)^48^ sustained VT was defined as run of consecutive ventricular beats (monomorphic or polymorphic) for greater than (for sustained) 30 seconds on telemetry or ECG, or termed non-sustained VT (NSVT) if less than 30 seconds of duration.^49^ CHB was defined as no evidence of atrioventricular contraction detected by ECG (3) at any time within follow-up after the first anthracycline dose.^50^

## Sample preparation and sequencing

### Analysis of genetic data

#### Quality control

Genotypes from exome sequencing as well as those from imputation were obtained from MER and Tapestry studies. Samples were evaluated for call rate, sex check, relatedness and population substructure utilizing biallelic variants having a genotyping rate ≥ 98%, minor allele frequency (MAF) ≥ 2%, and Hardy-Weinberg p-value > 5e-08 while excluding those in highly volatile regions of the genome using Plink.^51^ Samples with a call rate ≥ 98% and inbreeding coefficients on the X-chromosome ≤ 0.3 for females and 0.7 or greater for males were retained. Relatedness was evaluated using KING-robust^52^ implemented in Plink.^53^ Among samples that were related up to the third degree (kinship coefficient ≥ 0.0442), only one sample from each pair or family was retained. Population substructure was evaluated using Eigenstrat.^54^ Eigenstrat was set to remove population outliers beyond 6sd over five iterations.

Following QC procedures within each cohort, genotype data from both studies was combined to evaluate and identify relateds between MER and Tapestry. Utilizing a common set of variants, relatedness was evaluated using KING-robust in Plink v2 and related samples excluded, as described above.

### Imputation

#### Genome-wide association study and meta-analysis

Genotypes derived from imputed dosages of biallelic variants with an imputation R^2^≥0.9 and a MAF≥0.001 across all samples that remain after QC in MER and Tapestry studies were imported into Plink v2. Variants were tested for association with the binary phenotypes representing arrhythmia and heart failure with arrhythmia combined, (defined as “cardiac event”) using multivariate logistic regression analysis in Plink^53^ while adjusting for age at cancer diagnosis, biological sex, radiation therapy (yes=1/no=0) and the first three principal components adjusting for population substructure.

Summary statistics including chromosome, position, tested (A1) and untested (A2) alleles, odds ratios (OR) or beta coefficient (BETA), standard error, p-value and sample size of each tested variant for each phenotype in MER and Tapestry were exported to flat files. These were then utilized to perform meta-analysis in Plink v1.9 using the *‘--meta-analysis*’ function. Random-effects meta-analysis p-values and BETA/OR along with heterogeneity index (I^2^) and p-value for Cochran’s Q statistic evaluating heterogeneity between studies were utilized to interpret the findings.

To identify independent variants with the most significant association with a phenotype at any given locus, variants were pruned utilizing the “*--clump*” function in Plink v1.9. Clumping process groups variants with a linkage disequilibrium (ld) R^2^ greater than a given threshold and identifies index variant with the most significant p-value. Utilizing the MER imputed data and random effects meta-analysis p-values, variants in 50kb window (*--clump-kb 50*) were pruned to R^2^ of 0.2 (*--clump-r2 0.2*) to identify and retain index variants at each locus (reported in table 2 and supplementary table 1).

### In-Silico analysis

The transcription factor binding affinity and the fold difference in binding between the two DNA sequences for the *KCTD15* promoter variants, rs62101381 and rs141116709 were predicted *in silico* using the Transcription factor Affinity Prediction (TRAP) web tools,^55^ sTRAP difference between two sequences with matrix file transfac12.1, background model chordate conserved elements, and multiple test correction Benjamini-Hochberg.

#### Cardiomyocyte culture

AC16 Human Cardiomyocytes were purchased from the American Tissue Culture Centre (ATCC, USA). Cardiomyocytes were maintained in DMEM/F-12 (Dulbecco’s Modified Eagle Medium/Nutrient Mixture F-12) medium (ThermoFisher Cat# 11320033) supplemented with 12.5% Fetal Bovine Serum and 1% Penicillin-streptomycin. Cells were incubated at 37 °C in a humidified incubator containing 5% CO_2_. The media was changed every 2 days and cells were maintained for not more than 10 passages.

#### Quantitative PCR (qPCR) Gene expression

AC16 Human Cardiomyocytes were seeded at a density of 3 x 10^3^ cells per well in 6-well plates. After overnight incubation, the cells were treated with a vehicle control or 0.5 uM Doxorubicin for 48 hours (six replicates per group). Total RNA was extracted from the cardiomyocytes using RNeasy Mini Kit (Qiagen, USA) following the manufacturer’s instructions. RNA concentration and quality was determined using the NanoDrop 2000C spectrophotometer (ThermoFisher Scientific, USA) and the Agilent Bioanalyzer 2100 (Agilent Technologies, USA) respectively. High-Capacity cDNA Reverse Transcription Kit (ThermoFisher Cat# 4368813) was used to reverse transcribed each RNA sample into cDNA in a total volume of 20ul. Using the cDNA samples, gene expression was assessed by qPCR using Taqman Gene Expression Assay probes for MMP16 (Hs00234676_m1), KCTD15 (Hs00225337_m1), GAPDH (Hs99999905_m1) performed on a QuantStudio 3 Real-Time PCR System (Applied Biosystems, USA). Gene expression level of MMP16 and KCTD15 were assessed relative to GAPDH. Fold change was calculated using 2^−ΔΔCT^ formula.

### *In vivo* model of doxorubicin-induced cardiotoxicity

#### Animals

Zebrafish (Danio rerio) WIK strain were maintained under 14 hours light–10 hours dark cycle at 28.5°C and handled with care. The animal study protocols were approved by the Mayo Clinic Institutional Animal Care and Use Committee (IACUC) under protocol number A00002783-17.

#### Anthracycline-induced cardiotoxicity model in adult zebrafish

To establish a progressive anthracycline induced cardiotoxicity (AIC) model, we previously identified that a dose of 20 µg/g doxorubicin (Dox) represents the highest concentration that did not induce significant fish mortality while still leading to reduced cardiac function. This dose aligns closely with those commonly used in rodent AIC models (15–25 mg/kg)^56^ and approximates the cumulative dose threshold for cardiotoxicity in humans (550 mg/m², which is approximately equivalent to 15 mg/kg).^57^ In this manuscript, doxorubicin was administered via a single bolus of intraperitoneal (IP) injection, following the protocol previously described in our earlier publication. ^58^

#### MMEJ-based sgRNA design and F₀ genetic testing

Single guide RNA (sgRNA) design and F₀ embryo injections were carried out following a recently published MMEJ-based protocol.^59^ In brief, target exon sequences were submitted to the MENTHU web-based platform (http://genesculpt.org/menthu/) to identify microhomology-mediated end joining (MMEJ) target sites. sgRNA sequences with high MENTHU scores were selected for synthesis. Chemically modified sgRNAs were synthesized by Synthego (Synthego Corporation) and resuspended in nuclease-free duplex buffer (Integrated DNA Technologies, 11-01-03-01), then diluted to a final working concentration of 5 µM. To generate sgRNA–Cas9 ribonucleoprotein (sgRNP) complexes, sgRNAs were incubated with Cas9 protein and microinjected into one-cell stage zebrafish embryos. Injected F₀ embryos were individually collected for downstream analysis to assess knockout efficiency. Genomic DNA was extracted, and target regions were amplified and subjected to Sanger sequencing (Genewiz; https://clims4.genewiz.com/CustomerHome/Index). Resulting sequences were analyzed using the ICE v2 CRISPR Analysis Tool (Inference of CRISPR Edits; https://www.synthego.com/products/bioinformatics/crispr-analysis) to calculate knockdown efficiency scores.^60^ The primer sequences used to assess knockout efficiency were: mmp16a-MJ-F: GCATGTTTCAGATGCTCTGTCT R: AACTTCCTCAGCCCACAAGG mmp16b-MJ-F: TAGAGCCCATTGTGTGTATTCTG R: ACTTGTTTAGAGTCCCTGTGATG

### *In vivo* echocardiography for adult fish hearts

High-frequency echocardiography was performed at 28 days post injection (dpi) using a Vevo 3100 imaging system (Fujifilm VisualSonics Inc., Toronto, Canada) equipped with a 50-MHz linear array transducer (MX700) following previously established methods.^61^ Acoustic gel (Aquasonic 100, Parker Laboratories Inc.) was applied to the transducer surface, to ensure optimal coupling with the tissue interface. Zebrafish were anesthetized with tricaine (0.16 mg/mL) and positioned ventral side up on a soft-sponge stage. The MX700 transducer was aligned above the zebrafish to acquire sagittal imaging planes of the heart.^61^ Three to five independent cardiac cycles per fish were measured to determine the average values. Cardiac function indices of the ejection fraction (EF) was calculated using the formula: EF=(EDV−ESV)/EDV. EDV and ESV are the ventricular volumes at the end-diastolic stage and end-systolic stage, respectively.

## RESULTS

### Genetic data

Following comprehensive QC procedures, 2,494 participants in the MER study were retained after QC. Two samples with mismatched sex, 20 related and 76 population outliers were detected and removed. Similarly, in the Tapestry study, 1 sample was excluded for call rate, 1 for sex mismatch, 5 for relatedness and 52 were population outliers, leaving 1,049 after QC. Evaluation of relatedness between the two studies identified 16 pairs of related samples (twelve duplicates and four 1^st^ degree relatives). We excluded the 16 MER participants that were related to Tapestry from all downstream analysis, resulting in 2,478 post-QC participants in the MER and 1,049 participants in the Tapestry.

For samples that passed QC, genotypes imputed to the TOPMED R3 reference panel ^62^ using Minimac (v4.1.6) ^63^ and Eagle phasing (v2.4) ^64^ were obtained. Genotypes from imputation for variants with an R^2^ ≥ 0.9 and a MAF ≥ 0.001 were imported into Plink v2 for GWAS. Variants were annotated using ANNOVAR^65^.

### Patient characteristics

Following QC and ancestry analysis, the MER lymphoma dataset consisted of 1,261 patients with lymphoma, 77 of which had an outcome of arrhythmia and 1,184 patients who did not experience arrhythmia after anthracycline treatment (Table 1). The Tapestry dataset used for genetic analyses consisted of 125 patients with breast cancer and 54 patients with lymphoma, of which 13 patients had an outcome of arrhythmia and 172 patients who did not experience arrhythmia after anthracycline treatment (Table 1), yielding 90 unique arrhythmia cases (6.64%) and 1,356 patients with no arrhythmia (93.4%) tested in the meta-analysis.

**Table 1:** Patient Characteristics in MER and Tapestry GWAS. MER and Tapestry participants receiving anthracycline, number of arrhythmia cases and controls, with genotypic data after QC (and ancestry)

|  |  | MER dataset |  |  | Tapestry dataset |  |  |
| --- | --- | --- | --- | --- | --- | --- | --- |
|  |  | Arrhythmia | No arrhythmia | p-value | Arrhythmia | No arrhythmia | p-value |
| Baseline Characteristics |  |  |  |  |  |  |  |
| Cancer Type | Breast Cancer | 0 | 0 |  | 7 | 118 |  |
|  | Lymphoma | 77 | 1184 |  | 6 | 54 |  |
| Age | median (Range) | 65 (31 - 83) | 56 (18 - 93) | 2.00E-88 | 67 (43 - 89) | 55 (18 - 79) | 7.49E-03 |
| Sex | Female (%) | 21 (27) | 506 (43) | 0.029 | 7 (54) | 145 (84) | 1.02E-02 |
|  | Male (%) | 56 (73) | 678 (57) |  | 6 (46) | 27 (16) |  |
| BMI | n | 76 | 1160 | 0.46 | 13 | 171 | 0.11 |
|  | median (Range) | 27.4 (19 - 45.4) | 27.1 (16.9 - 49.6) |  | 30 (22 - 64.2) | 26.6 (18.8 - 51) |  |
| Diabetes | No (%) | 60 (78) | 953 (80) | 0.53 | 11 (85) | 159 (92) | 0.33 |
|  | Yes (%) | 6 (8) | 72 (6) |  | 2 (15) | 13 (8) |  |
|  | Missing (%) | 11 (14) | 159 (13) |  | 0 (0) | 0 (0) |  |
| Radiation Therapy | No (%) | 67 (87) | 991 (84) | 0.88 | 12 (92) | 166 (97) | 0.46 |
|  | Yes (%) | 10 (13) | 193 (16) |  | 1 (8) | 6 (3) |  |
| Outcomes |  |  |  |  |  |  |  |
| Any arrhythmia | N | 77 | 1184 |  | 13 | 172 |  |
| Atrial fibrillation | No |  |  |  | 3 | 172 | N/A |
|  | Yes |  |  |  | 10 | - |  |
| Ventricular tachycardia | No |  |  |  | 7 | 172 | N/A |
|  | Yes |  |  |  | 6 | - |  |
| Complete heart block | No |  |  |  | 12 | 172 | N/A |
|  | Yes |  |  |  | 1 | - |  |
| Heart Failure | No (%) | 55 (71) | 1158 (98) | 2.81E-19 | 12 (92) | 167 (97) | 0.37 |
|  | Yes (%) | 22 (29) | 26 (2) |  | 1 (8) | 5 (3) |  |

Type of arrhythmia was not collected in the MER study, but these data were available in Tapestry individuals, in which 10/13 patients experienced AF, 6/13 patients experienced VT and one patient experienced CHB attributed to doxorubicin. Four patients experienced more than one type of arrhythmia (at different time-points post-doxorubicin).

Body mass index measured (BMI), diabetes or radiation variables were not statistically different between patients with and without arrhythmia in either the MER or Tapestry (Table 1) datasets. We observed significant differences for older age and male sex in both datasets and these variables were incorporated into the final genetic model.

Not surprisingly, there was some overlap between arrhythmia and heart failure outcomes. In the larger MER dataset, 22/77 (28.6%) patients who experienced arrhythmia also experienced heart failure, compared to 26/1158 (2.2%) of patients that did not experience arrhythmia, p=2.81E-19. In the Tapestry dataset, the same trend was observed, 1/13 (7.7%) vs 5/172 (2.9%), but did not reach statistical significance, p=0.367.

### *CHST8:KCTD15* locus reached genome-wide significance for association with anthracycline-related arrhythmia

Meta-analysis of doxorubicin-related arrhythmia in the MER and Tapestry GWAS datasets (Figure 1A) identified a total of 98 independent variants, p<1×10^-5^, (Supplementary Table 1). The top independent loci, p<5E-06 are shown in Table 2, of which the most significant association by several orders of magnitude and reaching criteria for genome-wide significance mapped to the 3’ and 5’ flanking regions respectively of two genes, *CHST8* (carbohydrate sulfotransferase 8) and *KCTD15* (potassium channel tetramerization domain containing 15) on chromosome 19. Twelve variants at this locus were in complete linkage disequilibrium with p-values ranging 9.01×10^-9^ (sentinel variant, rs12151014) to 3.18×10^-8^ (Figure 1C, Supplementary Table 2).

**Figure 1:**
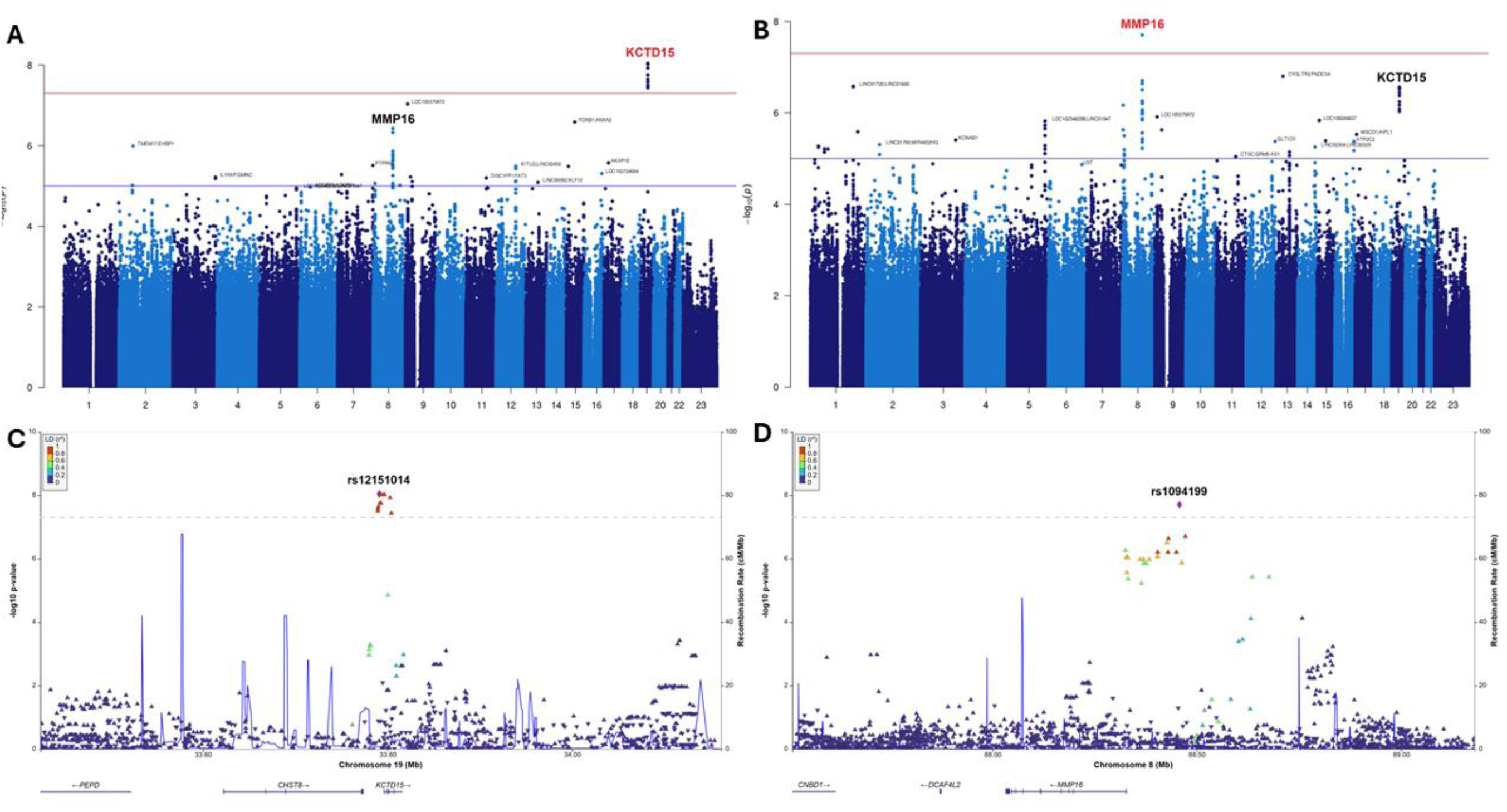
Meta-analysis of anthracycline-related GWAS datasets. A. Manhattan plot anthracycline-related arrhythmia; B. Manhattan plot anthracycline-related cardiac event (outcome of arrhythmia and/or heart failure); C. *CHST8*:*KCTD15* Locus Zoom plot anthracycline-related arrhythmia; D. *MMP16* Locus Zoom plot anthracycline-related cardiac event (outcome of arrhythmia and/or heart failure).

**Table 2:**
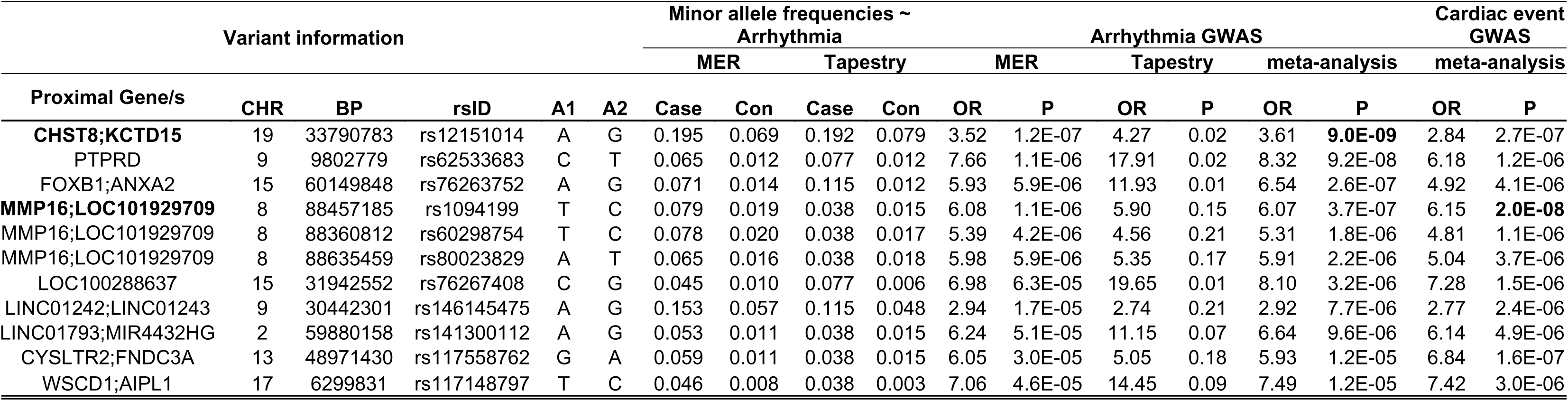
Variants associated with anthracycline-related arrhythmia and cardiac events at the chr 19 *CHST8*:*KCTD15 and chr 16 MMP16* loci. CHR, chromosome; BP, Base-pair coordinate Hg38, A1, Allele 1; A2, Allele 2; OR, Odds Ratio; Q, p-value for Cochran’s Q statistic; I, I^2^ heterogeneity index (0-100 scale). Allele freq, Allele frequency in cases with Arrhythmia (ARR) and without Arrhythmia (no ARR) in MER and Tapestry datasets. GWAS top hits for Arrhythmia and Cardiac Event (meta-analysis P < 5E-06 in either). Arrhythmia GWAS summary statistics from the MER and Tapestry studies as well as meta-analysis results for arrhythmia and cardiac event phenotypes. Independent variants (r2<0.2) with a meta-analysis p-value < 5E-06 in either outcome are tabulated. From left to right, gene or genes proximal to the variant, its chromosome, position, dbSNP ID (rsID), tested allele (A1), untested allele (A2), minor allele frequencies for arrhythmia cases and controls (Con) in MER and Tapestry, odds-ratio (OR) and p-values (P) for arrhythmia GWAS from MER, Tapestry and the combined meta-analysis as well as meta-analysis results for cardiac event are shown. Meta-analysis results (OR and P) for the random-effects model are shown. Test of heterogeneity (Cochran's Q statistic and I2 heterogeneity index) showed no significant heterogeneity between the cohorts in either meta-analyses (I=0, Q>0.4 in both for all variants shown). Proximal genes and meta-analysis P for the top variant in both the phenotypes are bolded. Cardiac event summary statistics from MER and Tapestry are not shown. I2 and Q values along with summary statistics for all variants with meta-analysis p<5E-05 for either phenotype are shown in Supplementary Table 1.

A breakdown of associated variants at the *CHST8*:*KCTD15* locus is further shown in (Supplementary Table 2). The sentinel variant, rs12151014, mapped 6.1 kb upstream of *KCTD15* and 17.3 kb downstream of *CSHT8*. The minor allele frequency in arrhythmia cases was 0.195 versus 0.069 in controls in the MER GWAS, OR = 3.52, p= 1.17 x 10^-7^ and 0.19 in cases versus 0.079 in controls in the Tapestry GWAS, OR=4.27, p= 0.024, increasing in significance to p = 9.01×10^-9^, OR 3.61 in the meta-analysis, Cochran’s Q = 0.78 and I^2^ = 0. 11. Other variants in R^2^ of 1 with the sentinel variant mapped to the *KCTD15* 5’ flanking region, of which rs62101381 and rs141116709 mapped within the promoter region, 798 and 523 bp upstream of exon 1 respectively.

### Multiple variants at the *MMP16* locus were suggestive of genome-wide association with anthracycline-related arrhythmia

Multiple independent variants at the *MMP16* (matrix metalloproteinase-16) locus on chromosome 8 were associated with doxorubicin-related arrhythmia in the meta-analysis (Table 2, Supplementary Table 1). This region included 24 variants (R^2^ with sentinel variant rs1094199, ranging 0.4-1.0), mapping to the *MMP16* 5’ flanking region, p-value ranging = 3.72×10^-7^ to 3.52×10^-5^ (Figure 1B,D, Supplementary Table 2).

The minor allele frequency of the sentinel variant in the arrhythmia cases was 0.079 versus 0.019 in controls in the MER GWAS, OR = 6.08, p = 1.09 x 10^-6^ and 0.038 in cases versus 0.015 in controls in the Tapestry GWAS, OR=5.90, p= 0.136, and p = 3.72 x 10^-7^, OR 6.07 in the meta-analysis, Cochran’s Q = 0.98 and I^2^ = 0, (Supplementary Table 2).

Other loci with suggestive genome-wide associations included *PTPRD* rs62533683, *FOXB1*;*ANXA2* rs76263752, *LOC100288637* rs76267408, *LINC01242*;*LINC01243* rs146145475, *LINC01793* rs141300112, *CYSLTR2*;*FNDC3A* rs117558762, *WSCD1*;*AIPL1* rs117148797 which were mostly single variant associations. Summary statistics of these variants are shown in Table 2 and Supplementary Table 1.

### Overlap of associated *CHST8*:*KCTD15* and *MMP16* variants with anthracycline-related heart failure

Our available sample size for heart failure as an outcome related to anthracycline in the MER and Tapestry datasets was limited. However, given the relationship between arrhythmia and development of heart failure and *vice versa*,^66,67^ we also explored the *CHST8*:*KCTD15* and *MMP16* loci for association with a combined analysis of heart failure and/or arrhythmia which we termed “cardiac event”. This analysis yielded a total number of 103 cases in the MER dataset and 18 cases in the Tapestry dataset. In this meta-analysis (Table 2), the top associated variant was at the *MMP*16 5’ flanking region, rs1094199, reaching genome-wide significance, p = 1.98 x 10^-8^, OR = 6.15. At the *CHST8*:*KCTD15* locus, the sentinel variant (rs12151014) remained the same as in the arrhythmia only analysis, but overall significance declined from p = 9.01 x 10^-9^, to p=2.73 x 10^-7^ and OR decreased from 3.61 to 2.84 (Figure 1B). QQ plots are shown in Supplementary Figure 2).

### No overlap of *MMP16*, *CHST8*:*KCTD15* arrhythmia-associated variants with Body Mass Index

We next assessed the *CHST8*:*KCTD15* and *MMP16* loci for genetic association with other known traits in the NHGRI-EBI Catalog of human genome-wide association studies.^68,69^ Neither locus is previously associated with arrhythmia, heart failure or cardiotoxicity in the GWAS Catalog, but both of these loci have significant associations with body mass index (BMI).^70–76^ Given the known correlation of arrhythmia and obesity in the literature^77–79^ and that our analyses did not include BMI as a covariate, we examined for overlap of associated variants, p < 1 x10^-5^ in this study with those associated with obesity or BMI in the GWAS catalog. No overlap was identified between *MMP16* and *CHST8*:*KCTD15* arrhythmia-associated variants and BMI-associated variants.

We also re-analyzed our arrhythmia GWAS in the MER dataset for association with BMI. None of the 24 variants at the *MMP16* locus, that were associated with arrhythmia, p<1×10^-5^ showed any association with BMI (p-values ranging 0.157 to 0.833). Similarly, at the *CHSTD8*:*KCTD15* locus, we did not observe any association (p<0.05) with BMI for variants that were associated with arrhythmia, p<1×10^-5^, suggesting that the observed associations with doxorubicin-related arrhythmia are independent of BMI.

### *KCTD15* and *MMP16,* but not *CHST8* are moderately expressed in human iPSC-derived cardiomyocytes

Gene expression in the heart was assessed for each locus of interest based on RNASeq analysis of human iPSC-derived cardiomyocytes from our previous publication.^80^ In human iPSC-derived cardiomyocytes, *KCTD15* and *MMP16* were present at moderate levels, with log counts per million (logCPM) of 3.38 and 4.88 respectively in a dataset ranging logCPM of -1.43 to 13.83. *CHST8* expression was not observed in this dataset, and in GTEx, *CHST8* is expressed most highly in pituitary tissue, followed by brain and breast mammary tissues.

Given the correlation of arrhythmia with aging,^81^ we also investigated differential gene expression of *CHST8*, *KCTD15* and *MMP16* in the hearts of aged macaque monkeys in a publicly available single cell sequencing dataset from the hearts of old and young cynomolgus macaque which annotated 18 different cardiac cell types.^82^ In the hearts of aged macaque monkeys, *MMP16* expression was significantly downregulated in cardiac endothelial cell types including arterial, capillary and vascular endothelial cells, with the largest fold change observed in arterial endothelial cells, log fold change = -0.875, adjusted p = 6.91 x10^-38^ but not in any other annotated cardiac cell type. Neither *CHST8* or *KCTD15* were differentially expressed in aged macaque heart for any cardiac cell type.

### *KCTD15* and *MMP16* gene expression in response to doxorubicin in AC16 cardiomyocytes

We next moved to assess *KCTD15* and *MMP16* gene expression in response to 0.5uM doxorubicin over 48 hours, using human AC16 cardiomyocytes. In this model, both *KCTD15* and *MMP16* expression was significantly reduced in response to doxorubicin, fold change 2.23, p<0.0001, Figure 2A and fold change 1.44, p=0.012, Figure 2B, respectively.

**Figure 2:**
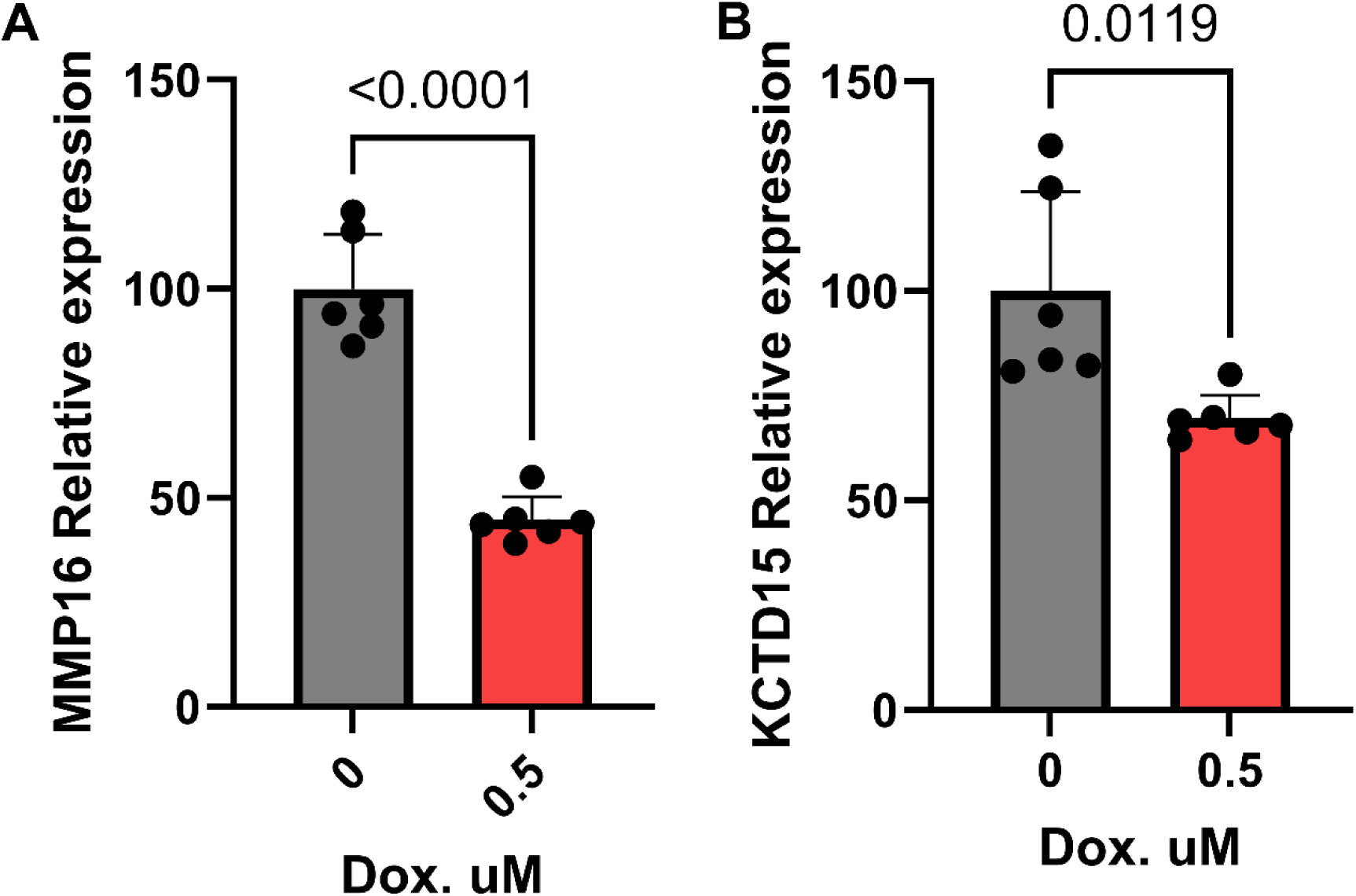
*MMP16* and *KCTD15* gene expression in AC16 cardiomyocytes in response to doxorubicin. *MMP16* (A) and KCTD15 (B) relative gene expression following 48 hours treatment with 0.5uM doxorubicin. Data are based on six replicates per group. Error bars are standard error of the mean (SEM).

### *In-silico* analysis of associated variants in the *KCTD15* promoter suggested differential binding of PAX9 and REST transcription factors to the risk and wild-type haplotypes

Two of the top associated variants at the *CHST8*:*KCTD15* locus (rs62101381 and rs141116709) mapped 275 bp apart, within the *KCTD15* promoter region (hg38:chr19+: 33,795,495-33,797,041) and were in complete linkage disequilibrium (R^2^=1). *In-silico* analysis of transcription factor binding affinity to the associated “risk” and “wild-type” haplotypes of these variants showed differential binding of the PAX9 transcription factor predicted to have significant binding affinity to the “wild-type sequence”, p=0.004 and not to the “risk sequence”, p=0.082, whereas, the NRSF (also known as REST) transcription was predicted to have significant binding affinity to the “risk” sequence, p=0.023 and not significantly to the “wild-type” sequence, p=0.10 (graphically shown in Supplementary Figure 3).

### *MMP16* CRISPR KO in F0 adult zebrafish demonstrated salutary modifying effects to doxorubicin-induced cardiomyopathy

Given the association between *MMP16* and arrhythmia and / or heart failure, we hypothesized that *MMP16* could play a role in AIC. To test this hypothesis, we adopted an adult zebrafish model of AIC for testing the potential modifying effect of a *mmp16* knockout. Due to the duplication of the zebrafish genome, two homologous isoforms of *mmp16, mmp16a* and *mmp16b*, were identified. We designed a sgRNAs for each isoform, which effectively knockout these isoforms at F0 generation, respectively (Figure 3A,B). In contrast to significantly reduced ejection fraction in wild type zebrafish treated with Dox (p<0.01), ejection fraction (EF%) was preserved at levels comparable to those of WT zebrafish without Dox treatment in *mmp16b* (Figure 3C), but not in *mmp16a* mutant fish. These genetic data in zebrafish suggested *mmp16* as a salutary modifier for AIC, and *mmp16b* as the predominant functional isoform of *mmp16* in a zebrafish heart.

**Figure 3:**
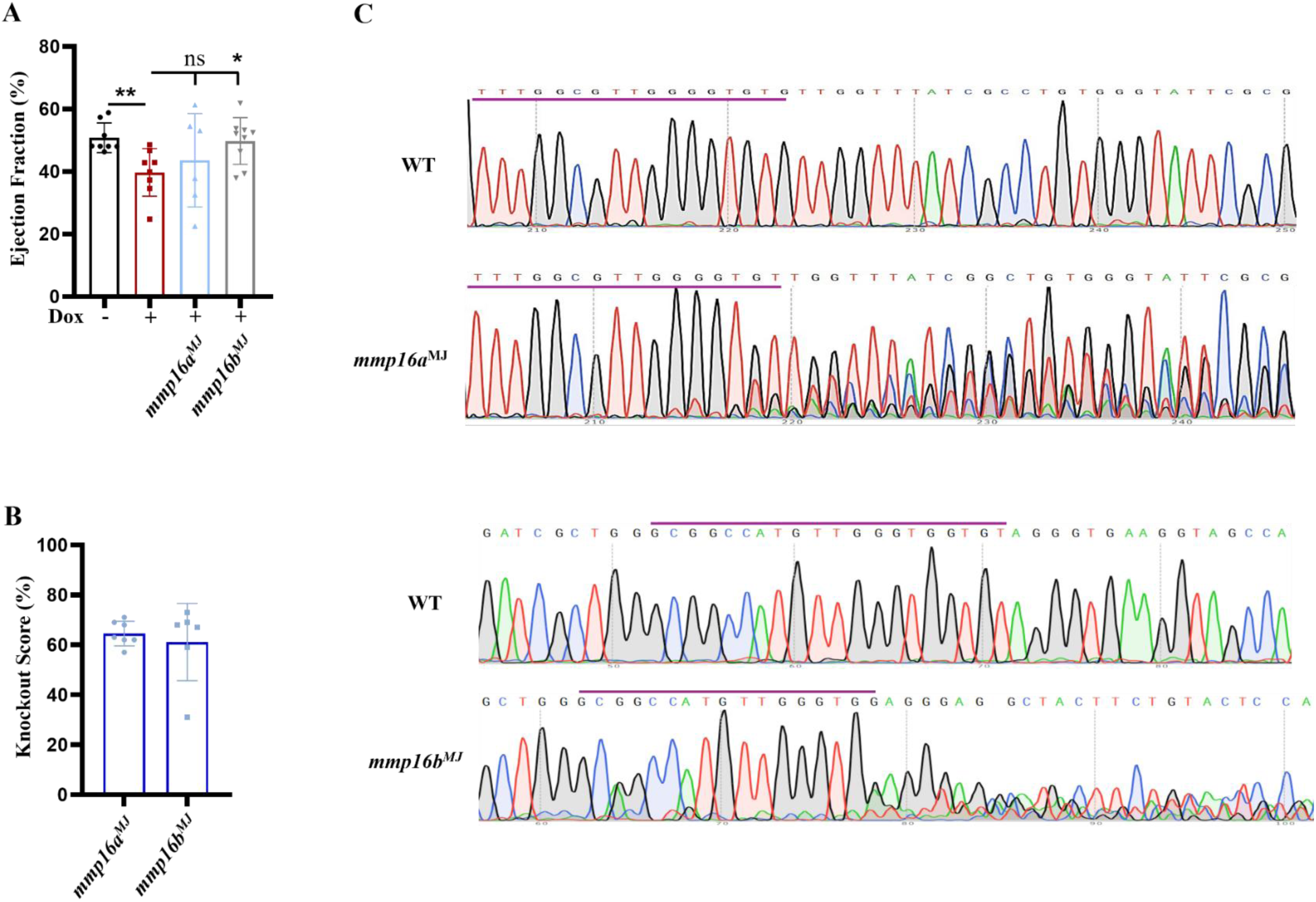
*mmp16* knockout preserved ejection fraction in adult zebrafish model of doxorubicin-induced cardiomyopathy. (A) Ejection fraction in wild-type zebrafish +/-doxorubicin and in *mmp16a* and *mmp16b* knockout fish treated with doxorubicin. *p<0.05, **p<0.01, ns, not significant. (B) *mmp16a* (mmp16a-MJ) and *mmp16b* (mmp16b-MJ) Crispr KO scores. of (C) Sanger sequencing wild-type (WT) *mmp16* and knockout of *mmp16a* (mmp16a-MJ) and *mmp16b* (mmp16b-MJ).

## Discussion

Defining novel risk genes and variants for anthracycline-induced cardiac outcomes has clinical importance towards development of personalized medicine approaches. Lack of replication due to the “winners curse” ^83^ is a common problem with the traditional approach of discovery cohort followed by replication in a second (often smaller available sample). We chose a different approach, taking two very similar (genetically homogeneous populations) straight to meta-analysis, on the basis that any genome-wide significant hits (p<5×10^-8^) would be worthy of further phenotypic exploration and functional validation. We used doxorubicin-related arrhythmia (as it was more prevalent than heart failure) as the primary outcome, and further explored the phenotype by including heart failure in the outcome.

Two novel loci were identified, *CHST8*:*KCTD15* and *MMP16*, as associated with arrhythmia or anthracycline-related cardiac outcomes. Neither of these have been previously described with this association, likely because we focused primarily on arrhythmia and the majority of genetic association studies of doxorubicin-related cardiotoxicity have used binary outcomes of LVEF decline by at least 10% points to <50,^84^ symptomatic heart failure^85,86^ or quantitative analysis of the maximum decline in LVEF.^87,88^ To our knowledge, this is the first genetic study of anthracycline-related arrhythmia. Further analyses of the CHST8:KCTD15 locus focused on KCTD15, because CHST8 is not expressed in cardiomyocytes or other cardiac tissues, and the sentinel variant mapped to the KCTD15 promoter region with suggestive in silico evidence of the sentinel variant increasing the binding affinity of a transcriptional repressor that is important to cardiac gene expression, discussed below.

*KCTD15* belongs to the family of potassium channel tetramerization domain (KCTD) containing proteins, which includes a total of 26 members that share sequence similarity with the cytoplasmic domain of voltage-gated K+ channels.^89^ *KCTD15* is ubiquitously expressed in the body, with moderate expression in left ventricular heart tissue (GTEx) and moderate to high expression in our previously published transcriptome dataset of human iPSC-derived cardiomyocytes.^90,91^ Little is known about the biological function of *KCTD15* in normal physiologic conditions, but it is known to be overexpressed in acute lymphoid and acute myeloid leukemia^92^ and in HER2+ breast cancer,^93^ and in the SKBR3 HER2+ cell line, *KCTD15* knock-out significantly reduced cell proliferation, cell cycle progression and increased sensitivity to doxorubicin.^93^ In our study, the genetic variants most strongly associated with doxorubicin-related arrhythmia, map to the *KCTD15* promoter region, for which *in-silico* analysis suggests could alter the binding affinity of NRSF/REST (neuron-restrictive silencer factor), a transcriptional repressor, important for regulation of the fetal cardiac gene program.^94^

Mouse models with dominant negative NRSF demonstrate several abnormalities in cardiac electrophysiological properties and ion channel expression^95,96^ and these abnormalities were ameliorated by inhibition of N-type calcium channels, which prevented lethal arrhythmia’s in mice with heart failure.^97^ Replication of the *KCTD15* genetic associations and physical validation of the relationship between these variants and NRSF binding are required, but we consider these findings as preliminary data to postulate the hypothesis that genetic variants at the *KCTD15* promoter identify a sub-population of patients at risk for doxorubicin-induced arrhythmia, and inhibition of KCTD15 or N-type calcium channels could be a precision therapy for this group of patients. Given that *KCTD15* silencing in the tumor reduced cell proliferation and increased sensitivity to doxorubicin, inhibition of KCTD15 could also be antineoplastic, allowing lower, less toxic dosing of doxorubicin.

Regarding genetic associations of *KCTD15* with other phenotypic outcomes, this locus is strongly associated with BMI in the GWAS catalog and there is a relationship between obesity and risk of arrhythmia.^77–79^ We emphasize that we did not observe association between arrhythmia cases and controls in either of our GWAS datasets and the associated variants with doxorubicin-related arrhythmia did not overlap with other GWAS hits for obesity and we conclude the arrhythmia association is independent of obesity.

The second locus identified in the primary meta-analysis of doxorubicin-related arrhythmia, was *MMP16*, a member of the matrix metalloproteinase family, a group of over 20 genes with biological roles in extracellular matrix remodeling, cardiovascular disease (particularly atrial remodeling in atrial fibrillation^98–104^) and cancer.^105^ In mouse models of doxorubicin-induced cardiotoxicity, oral inhibitors of MMP-2 attenuated doxorubicin-induced systolic and diastolic dysfunction^106^ and preventing extracellular matrix remodeling.^107^

Less is known of the relationship of *MMP16* and cardiotoxicity, but there is some evidence of a role for MMP16 in the biology of atrial fibrillation and cancer, whereby the ratio of MMP16 and its inhibitor, TIMP4, was significantly decreased in atrial fibrillation patients with severe aortic stenosis^98^ and inhibition of MMP16 via MicroRNA-377-3p inhibited ovarian cancer cell growth, invasion and interstitial transition.^108^ In our study, we observed a significant decrease in *MMP16* expression in human cardiomyocytes (AC16) treated with doxorubicin and in our *in vivo* model of doxorubicin-induced heart failure, we found that *MMP16* knockout ameliorated the doxorubicin-induced decline in LVEF. Taken together, these data suggest that a sub-population of patients with *MMP16* genetic variants are susceptible to doxorubicin-related cardiac remodeling, which could potentially be prevented by pharmacological inhibition of MMP16, and perhaps that MMP16 inhibition may have both cardioprotective and anti-neoplastic capabilities.

The main caveats of the study are the relatively small sample size, lack of functional validation at the level of the variant (gene level only) and *in vitro* and *in vivo* models did not include assessment of arrhythmia, which was the primary outcome of our genetic screening in humans. Finally, although our current data imply a role of MMP16 and KCTD15 in cardiotoxicity, our studies did not elucidate the mechanism, which we consider a future direction. Nonetheless, our study found genome-wide significance of doxorubicin-related arrhythmia in two genetically homogenous patient populations at two novel loci, *KCTD15* and *MMP16*. We demonstrated *in-silico* and/or functional validation of their roles in response to doxorubicin and as cardioprotective therapeutic targets, supported by evidence from the literature for these genes as potential therapeutic targets in cancer.

The identification of novel genetic loci associated with anthracycline related arrhythmia carries significant clinical implications. By detecting CHST8:KCTD15 and MMP16 as potential risk loci, this study provides a foundation for the development of predictive genetic screening tools that could stratify patients by susceptibility to cardiotoxicity prior to chemotherapy initiation. Such risk stratification may allow clinicians to tailor cancer therapy regimens, balancing therapeutic efficacy with cardiovascular safety. High-risk individuals could benefit from enhanced cardiac monitoring, early initiation of cardioprotective therapies, or consideration of alternative, less cardiotoxic agents. As such, understanding the genetic associations of anthracycline-induced arrhythmia may enable mechanistic studies that could inform novel therapeutic targets or preventive strategies. As precision oncology advances, integrating genetic risk factors into cardio-oncology protocols has the potential to significantly reduce treatment-related morbidity and improve long-term outcomes in cancer survivors.

---

**Supplementary Table 1:** GWAS top hits for Arrhythmia and Cardiac Event (meta-analysis P < 5E-05 in either). Arrhythmia GWAS summary statistics from the MER and Tapestry studies as well as meta-analysis results for arrhythmia and cardiac event phenotypes. Only independent variants (r2<0.2) with a meta-analysis p-value < 5e-05 in either phenotypes are tabulated. From left to right, gene or genes proximal to the variant, its chromosome, position, dbSNP ID (rsID), tested allele (A1), untested allele (A2), minor allele frequencies for arrhythmia cases and controls in MER and Tapestry, odds-ratio (OR) and p-values (P) for arrhythmia GWAS from MER, Tapestry and the combined meta-analysis as well as meta-analysis results for cardiac event are shown. Cochran’s Q statistic and heterogeneity index (I) evaluating heterogeneity between the two cohorts are provided along with OR and P for the random-effects model. Proximal genes and meta-analysis P for the top variant in both the phenotypes are bolded. Cardiac event summary statistics from MER and Tapestry are not shown.

| Variant information |  |  |  |  |  | Minor allele frequencies ~ Arrhythmia |  |  |  | Arrhythmia GWAS |  |  |  |  |  |  |  | Cardiac event GWAS |  |  |  |
| --- | --- | --- | --- | --- | --- | --- | --- | --- | --- | --- | --- | --- | --- | --- | --- | --- | --- | --- | --- | --- | --- |
|  |  |  |  |  |  | MER |  | Tapestry |  | MER |  | Tapestry |  | Meta-analysis |  |  |  | Meta-analysis |  |  |  |
| Proximal Gene/s | CHR | BP | rsID | A1 | A2 | Cases | Controls | Cases | Controls | OR | P | OR | P | OR | P | Q | I | OR | P | Q | I |
| CHST8;KCTD15 | 19 | 33790783 | rs12151014 | A | G | 0.195 | 0.069 | 0.192 | 0.079 | 3.52 | 1.2E-07 | 4.27 | 2.4E-02 | 3.61 | **9.0E-09** | 0.78 | 0 | 2.84 | 2.7E-07 | 0.63 | 0 |
| PTPRD | 9 | 9802779 | rs62533683 | C | T | 0.065 | 0.012 | 0.077 | 0.012 | 7.66 | 1.1E-06 | 17.91 | 2.3E-02 | 8.32 | 9.2E-08 | 0.52 | 0 | 6.18 | 1.2E-06 | 0.81 | 0 |
| FOXB1;ANXA2 | 15 | 60149848 | rs76263752 | A | G | 0.071 | 0.014 | 0.115 | 0.012 | 5.93 | 5.9E-06 | 11.93 | 1.1E-02 | 6.54 | 2.6E-07 | 0.51 | 0 | 4.92 | 4.1E-06 | 0.67 | 0 |
| MMP16;LOC101929709 | 8 | 88457185 | rs1094199 | T | C | 0.079 | 0.019 | 0.038 | 0.015 | 6.08 | 1.1E-06 | 5.90 | 1.5E-01 | 6.07 | 3.7E-07 | 0.98 | 0 | 6.15 | **2.0E-08** | 0.52 | 0 |
| TMEM17;EHBP1 | 2 | 62607935 | rs144003047 | A | G | 0.267 | 0.144 | 0.250 | 0.182 | 2.59 | 3.4E-06 | 2.60 | 1.3E-01 | 2.59 | 1.0E-06 | 1.00 | 0 | 1.99 | 9.1E-05 | 0.48 | 0 |
| MMP16;LOC101929709 | 8 | 88360812 | rs60298754 | T | C | 0.078 | 0.020 | 0.038 | 0.017 | 5.39 | 4.2E-06 | 4.56 | 2.1E-01 | 5.31 | 1.8E-06 | 0.89 | 0 | 4.81 | 1.1E-06 | 0.45 | 0 |
| MMP16;LOC101929709 | 8 | 88635459 | rs80023829 | A | T | 0.065 | 0.016 | 0.038 | 0.018 | 5.98 | 5.9E-06 | 5.35 | 1.7E-01 | 5.91 | 2.2E-06 | 0.93 | 0 | 5.04 | 3.7E-06 | 0.63 | 0 |
| AKAP10 | 17 | 19930961 | rs139899843 | A | G | 0.052 | 0.010 | 0.077 | 0.006 | 6.62 | 3.8E-05 | 23.71 | 1.5E-02 | 7.63 | 2.7E-06 | 0.35 | 0 | 5.79 | 1.7E-05 | 0.53 | 0 |
| PTPRN2 | 7 | 157695654 | rs1628432 | T | C | 0.084 | 0.025 | 0.077 | 0.026 | 4.75 | 1.2E-05 | 4.84 | 1.0E-01 | 4.76 | 3.1E-06 | 0.99 | 0 | 3.20 | 2.7E-04 | 0.90 | 0 |
| KITLG;LINC02458 | 12 | 88892039 | rs116953322 | G | A | 0.097 | 0.025 | 0.038 | 0.015 | 4.44 | 4.8E-06 | 3.22 | 3.6E-01 | 4.35 | 3.2E-06 | 0.81 | 0 | 3.16 | 1.2E-04 | 0.60 | 0 |
| LOC100288637 | 15 | 31942552 | rs76267408 | C | G | 0.045 | 0.010 | 0.077 | 0.006 | 6.98 | 6.3E-05 | 19.65 | 1.2E-02 | 8.10 | 3.2E-06 | 0.42 | 0 | 7.28 | 1.5E-06 | 0.72 | 0 |
| LOC102724084 | 16 | 80261322 | rs62049538 | A | G | 0.045 | 0.006 | 0.038 | 0.015 | 11.50 | 5.4E-06 | 3.39 | 3.1E-01 | 9.38 | 4.9E-06 | 0.35 | 0 | 7.13 | 1.6E-05 | 0.84 | 0 |
| AGR3;AHR | 7 | 17044830 | rs73680144 | A | T | 0.162 | 0.069 | 0.167 | 0.080 | 2.72 | 5.4E-05 | 5.79 | 2.0E-02 | 2.93 | 5.2E-06 | 0.34 | 0 | 2.16 | 4.2E-04 | 0.80 | 0 |
| LINC01239;LOC101929563 | 9 | 23077093 | rs149808175 | T | G | 0.052 | 0.014 | 0.077 | 0.018 | 5.42 | 1.4E-04 | 13.06 | 9.7E-03 | 6.28 | 5.9E-06 | 0.42 | 0 | 5.89 | 9.2E-04 | 0.22 | 33 |
| IL1RAP;GMNC | 3 | 190702364 | rs76730956 | T | C | 0.039 | 0.005 | 0.038 | 0.012 | 11.26 | 2.3E-05 | 7.81 | 1.0E-01 | 10.57 | 6.0E-06 | 0.79 | 0 | 7.02 | 1.3E-04 | 0.54 | 0 |
| DISC1FP1;FAT3 | 11 | 91465383 | rs111920676 | G | A | 0.058 | 0.015 | 0.038 | 0.012 | 5.89 | 2.3E-05 | 7.68 | 1.1E-01 | 6.04 | 6.3E-06 | 0.84 | 0 | 3.80 | 5.5E-04 | 0.97 | 0 |
| KITLG;LINC02458 | 12 | 88759339 | rs12300733 | C | A | 0.091 | 0.024 | 0.038 | 0.017 | 4.70 | 6.7E-06 | 1.89 | 6.1E-01 | 4.40 | 7.6E-06 | 0.48 | 0 | 2.87 | 9.8E-04 | 0.41 | 0 |
| LINC01242;LINC01243 | 9 | 30442301 | rs146145475 | A | G | 0.153 | 0.057 | 0.115 | 0.048 | 2.94 | 1.7E-05 | 2.74 | 2.1E-01 | 2.92 | 7.7E-06 | 0.93 | 0 | 2.77 | 2.4E-06 | 0.45 | 0 |
| LINC00392;KLF12 | 13 | 73589093 | rs17090303 | A | G | 0.032 | 0.005 | 0.038 | 0.006 | 11.62 | 6.4E-05 | 19.65 | 4.5E-02 | 12.55 | 8.1E-06 | 0.74 | 0 | 8.06 | 1.4E-04 | 0.90 | 0 |
| FAM86B3P;PRAG1 | 8 | 8296405 | rs17155266 | T | A | 0.045 | 0.009 | 0.038 | 0.006 | 6.68 | 8.4E-05 | 21.26 | 2.6E-02 | 7.59 | 8.6E-06 | 0.43 | 0 | 5.77 | 4.6E-05 | 0.70 | 0 |
| LINC01793;MIR4432HG | 2 | 59880158 | rs141300112 | A | G | 0.053 | 0.011 | 0.038 | 0.015 | 6.24 | 5.1E-05 | 11.15 | 6.7E-02 | 6.64 | 9.6E-06 | 0.68 | 0 | 6.14 | 4.9E-06 | 0.97 | 0 |
| COL14A1 | 8 | 120179902 | rs144842503 | A | G | 0.032 | 0.005 | 0.038 | 0.009 | 11.43 | 6.0E-05 | 12.46 | 6.2E-02 | 11.60 | 9.6E-06 | 0.95 | 0 | 7.60 | 1.7E-04 | 0.87 | 0 |
| LINGO2;LINC01242 | 9 | 30280270 | rs147145745 | T | C | 0.074 | 0.018 | 0.077 | 0.024 | 4.78 | 4.0E-05 | 5.67 | 1.0E-01 | 4.87 | 1.0E-05 | 0.88 | 0 | 3.69 | 1.3E-04 | 0.81 | 0 |
| ADGRF5;ADGRF1 | 6 | 46989654 | rs62401797 | G | C | 0.153 | 0.060 | 0.154 | 0.074 | 2.99 | 3.0E-05 | 2.70 | 1.6E-01 | 2.95 | 1.1E-05 | 0.90 | 0 | 2.29 | 2.3E-04 | 0.81 | 0 |
| LINC02737;CNTN5 | 11 | 97539172 | rs147540700 | A | G | 0.032 | 0.004 | 0.038 | 0.006 | 10.42 | 1.0E-04 | 20.38 | 3.7E-02 | 11.51 | 1.1E-05 | 0.67 | 0 | 7.07 | 2.9E-04 | 0.94 | 0 |
| CEP295 | 11 | 93688335 | rs117785479 | G | A | 0.045 | 0.008 | 0.038 | 0.009 | 8.58 | 2.3E-05 | 5.30 | 2.3E-01 | 8.11 | 1.1E-05 | 0.75 | 0 | 5.48 | 2.4E-04 | 0.82 | 0 |
| CYSLTR2;FNDC3A | 13 | 48971430 | rs117558762 | G | A | 0.059 | 0.011 | 0.038 | 0.015 | 6.05 | 3.0E-05 | 5.05 | 1.8E-01 | 5.93 | 1.2E-05 | 0.89 | 0 | 6.84 | 1.6E-07 | 0.84 | 0 |
| WSCD1;AIPL1 | 17 | 6299831 | rs117148797 | T | C | 0.046 | 0.008 | 0.038 | 0.003 | 7.06 | 4.6E-05 | 14.45 | 9.4E-02 | 7.49 | 1.2E-05 | 0.67 | 0 | 7.42 | 3.0E-06 | 0.83 | 0 |
| DISC1FP1;FAT3 | 11 | 91533238 | rs11019511 | A | G | 0.058 | 0.016 | 0.038 | 0.015 | 5.67 | 3.1E-05 | 5.40 | 1.7E-01 | 5.64 | 1.2E-05 | 0.97 | 0 | 3.48 | 1.1E-03 | 0.74 | 0 |
| KITLG;LINC02458 | 12 | 88944835 | rs74813500 | G | A | 0.092 | 0.025 | 0.038 | 0.015 | 4.14 | 1.8E-05 | 3.26 | 3.5E-01 | 4.08 | 1.2E-05 | 0.86 | 0 | 2.99 | 3.4E-04 | 0.64 | 0 |
| LINC01793;MIR4432HG | 2 | 59824634 | rs182866938 | T | A | 0.053 | 0.011 | 0.038 | 0.018 | 6.01 | 6.8E-05 | 10.50 | 7.2E-02 | 6.38 | 1.3E-05 | 0.69 | 0 | 5.02 | 6.5E-05 | 0.94 | 0 |
| AMZ1 | 7 | 2688578 | rs887452 | T | C | 0.058 | 0.015 | 0.083 | 0.018 | 4.99 | 1.1E-04 | 6.63 | 4.7E-02 | 5.22 | 1.4E-05 | 0.78 | 0 | 3.52 | 6.5E-04 | 0.85 | 0 |
| LY86-AS1 | 6 | 6613844 | rs373941465 | T | TC | 0.071 | 0.017 | 0.038 | 0.018 | 5.35 | 1.2E-05 | 1.91 | 6.1E-01 | 4.91 | 1.5E-05 | 0.43 | 0 | 3.32 | 6.3E-04 | 0.32 | 0 |
| LINC01505;ZNF462 | 9 | 106697591 | rs74552897 | A | G | 0.071 | 0.019 | 0.038 | 0.020 | 4.96 | 3.0E-05 | 3.92 | 2.6E-01 | 4.86 | 1.6E-05 | 0.85 | 0 | 3.08 | 1.5E-03 | 0.71 | 0 |
| FHIT | 3 | 60957538 | rs200466589 | A | ATCT | 0.322 | 0.183 | 0.231 | 0.175 | 2.16 | 4.3E-05 | 2.39 | 1.7E-01 | 2.18 | 1.6E-05 | 0.88 | 0 | 1.72 | 8.7E-04 | 0.47 | 0 |
| PRAG1 | 8 | 8325886 | rs2203826 | C | T | 0.112 | 0.042 | 0.154 | 0.035 | 3.06 | 1.5E-04 | 5.60 | 3.1E-02 | 3.29 | 1.7E-05 | 0.48 | 0 | 2.86 | 3.0E-05 | 0.84 | 0 |
| SEMA6A | 5 | 116478466 | rs150918584 | G | A | 0.039 | 0.008 | 0.038 | 0.006 | 7.05 | 1.7E-04 | 33.02 | 2.2E-02 | 8.29 | 1.7E-05 | 0.34 | 0 | 5.36 | 4.7E-04 | 0.45 | 0 |
| PDZD2 | 5 | 31734565 | rs28541418 | T | C | 0.079 | 0.020 | 0.038 | 0.012 | 4.32 | 6.5E-05 | 7.95 | 1.0E-01 | 4.52 | 1.8E-05 | 0.65 | 0 | 3.32 | 3.4E-04 | 0.98 | 0 |
| RERE | 1 | 8455558 | rs143104616 | G | A | 0.039 | 0.006 | 0.038 | 0.003 | 8.68 | 7.8E-05 | 55.81 | 7.0E-02 | 9.65 | 2.0E-05 | 0.42 | 0 | 6.87 | 6.0E-04 | 0.31 | 3 |
| LINC00477;BCAT1 | 12 | 24706894 | rs35484009 | A | G | 0.104 | 0.036 | 0.077 | 0.026 | 3.52 | 7.2E-05 | 4.46 | 1.2E-01 | 3.60 | 2.1E-05 | 0.82 | 0 | 2.75 | 3.1E-04 | 0.92 | 0 |
| ZBTB20 | 3 | 114817488 | rs112613950 | G | A | 0.104 | 0.035 | 0.125 | 0.068 | 3.63 | 2.6E-05 | 2.01 | 3.4E-01 | 3.32 | 2.1E-05 | 0.45 | 0 | 2.24 | 2.9E-03 | 0.44 | 0 |
| LARP4 | 12 | 50459878 | rs561048841 | G | C | 0.026 | 0.003 | 0.038 | 0.003 | 12.58 | 1.4E-04 | 19.72 | 5.8E-02 | 13.47 | 2.2E-05 | 0.79 | 0 | 8.82 | 2.5E-04 | 0.81 | 0 |
| LIMA1 | 12 | 50211382 | rs192444952 | A | G | 0.026 | 0.003 | 0.038 | 0.003 | 12.55 | 1.4E-04 | 19.68 | 5.8E-02 | 13.44 | 2.2E-05 | 0.79 | 0 | 8.80 | 2.6E-04 | 0.81 | 0 |
| EPC2;KIF5C | 2 | 148807313 | rs116474328 | G | A | 0.078 | 0.022 | 0.038 | 0.009 | 5.05 | 1.4E-05 | 1.20 | 9.0E-01 | 4.62 | 2.2E-05 | 0.34 | 0 | 4.08 | 2.7E-05 | 0.35 | 0 |
| GABRA4;GABRB1 | 4 | 47014242 | rs13139454 | C | G | 0.408 | 0.262 | 0.500 | 0.296 | 1.99 | 1.2E-04 | 2.38 | 6.8E-02 | 2.04 | 2.3E-05 | 0.72 | 0 | 1.91 | 2.8E-03 | 0.24 | 28 |
| CPHXL;CNTNAP4 | 16 | 75914587 | rs79148027 | G | A | 0.079 | 0.023 | 0.077 | 0.018 | 3.74 | 1.9E-04 | 9.86 | 2.8E-02 | 4.14 | 2.3E-05 | 0.38 | 0 | 2.69 | 2.2E-03 | 0.63 | 0 |
| TSNARE1 | 8 | 142304996 | rs74950006 | A | G | 0.026 | 0.003 | 0.038 | 0.003 | 13.39 | 2.1E-04 | 22.21 | 4.0E-02 | 14.64 | 2.3E-05 | 0.76 | 0 | 9.12 | 3.4E-04 | 0.94 | 0 |
| GAK | 4 | 857451 | rs193280704 | C | T | 0.019 | 0.003 | 0.077 | 0.003 | 13.33 | 8.8E-04 | 38.18 | 7.0E-03 | 17.33 | 2.4E-05 | 0.50 | 0 | 11.12 | 2.8E-04 | 0.54 | 0 |
| DNAJC11;LINC01672 | 1 | 6719318 | rs116724358 | G | A | 0.060 | 0.015 | 0.038 | 0.009 | 5.21 | 6.5E-05 | 6.26 | 1.7E-01 | 5.30 | 2.4E-05 | 0.89 | 0 | 3.55 | 8.5E-04 | 0.86 | 0 |
| ACACA | 17 | 37343847 | rs117651221 | A | G | 0.097 | 0.037 | 0.038 | 0.021 | 3.66 | 4.0E-05 | 3.21 | 3.2E-01 | 3.62 | 2.4E-05 | 0.91 | 0 | 2.55 | 1.2E-03 | 0.68 | 0 |
| LOC349160;PTN | 7 | 137220274 | rs186422040 | A | G | 0.032 | 0.003 | 0.038 | 0.009 | 11.29 | 1.2E-04 | 10.46 | 8.2E-02 | 11.14 | 2.4E-05 | 0.96 | 0 | 7.64 | 3.0E-04 | 0.69 | 0 |
| PTPRD-AS2;TYRP1 | 9 | 11158911 | rs1931426 | A | C | 0.208 | 0.104 | 0.154 | 0.070 | 2.34 | 1.3E-04 | 3.03 | 6.8E-02 | 2.41 | 2.5E-05 | 0.69 | 0 | 2.07 | 1.2E-04 | 0.79 | 0 |
| LOC107985115;NLRP3 | 1 | 247414803 | rs77563288 | A | T | 0.052 | 0.011 | 0.038 | 0.020 | 6.64 | 3.1E-05 | 2.73 | 3.3E-01 | 5.76 | 2.6E-05 | 0.43 | 0 | 3.73 | 1.2E-03 | 0.42 | 0 |
| OLIG3;LOC102723649 | 6 | 137633202 | rs12199435 | A | G | 0.250 | 0.133 | 0.154 | 0.122 | 2.42 | 4.4E-05 | 1.83 | 2.8E-01 | 2.33 | 2.7E-05 | 0.64 | 0 | 2.02 | 7.9E-05 | 0.42 | 0 |
| ADAMTS17 | 15 | 100318850 | rs78119169 | A | G | 0.052 | 0.008 | 0.038 | 0.006 | 7.01 | 3.1E-05 | 3.07 | 4.7E-01 | 6.55 | 2.7E-05 | 0.61 | 0 | 4.56 | 4.7E-04 | 0.54 | 0 |
| LINC02605;MIR12123 | 8 | 79211537 | rs6473143 | C | T | 0.169 | 0.074 | 0.154 | 0.073 | 2.62 | 7.8E-05 | 2.61 | 1.6E-01 | 2.62 | 2.8E-05 | 1.00 | 0 | 2.03 | 8.0E-04 | 0.56 | 0 |
| NRG3;LINC02650 | 10 | 83547388 | rs140352242 | A | G | 0.091 | 0.033 | 0.077 | 0.017 | 3.46 | 1.5E-04 | 5.94 | 6.5E-02 | 3.65 | 2.8E-05 | 0.59 | 0 | 2.93 | 1.9E-04 | 0.79 | 0 |
| EFNA3;EFNA1 | 1 | 155124619 | rs111742972 | A | C | 0.072 | 0.023 | 0.077 | 0.020 | 4.05 | 1.7E-04 | 6.09 | 6.1E-02 | 4.27 | 2.9E-05 | 0.69 | 0 | 2.88 | 1.5E-03 | 0.73 | 0 |
| DAB2IP | 9 | 121631358 | rs117843215 | T | C | 0.039 | 0.006 | 0.038 | 0.006 | 7.53 | 2.1E-04 | 17.56 | 4.5E-02 | 8.38 | 3.0E-05 | 0.58 | 0 | 6.74 | 7.7E-05 | 0.94 | 0 |
| LOC102724849 | 2 | 215459929 | rs114212774 | C | T | 0.033 | 0.005 | 0.038 | 0.003 | 9.66 | 1.3E-04 | 16.16 | 9.3E-02 | 10.24 | 3.1E-05 | 0.77 | 0 | 6.63 | 5.5E-04 | 0.74 | 0 |
| MMP8 | 11 | 102718321 | rs34829643 | T | C | 0.045 | 0.009 | 0.115 | 0.020 | 6.21 | 1.9E-04 | 5.39 | 6.4E-02 | 6.02 | 3.1E-05 | 0.89 | 0 | 4.59 | 1.7E-04 | 0.88 | 0 |
| MIR4471;ANKRD46 | 8 | 100387541 | rs145454677 | T | C | 0.058 | 0.014 | 0.077 | 0.012 | 4.47 | 3.3E-04 | 13.76 | 2.1E-02 | 5.11 | 3.2E-05 | 0.35 | 0 | 4.00 | 2.0E-04 | 0.43 | 0 |
| SDR16C5 | 8 | 56307347 | rs117160043 | G | T | 0.079 | 0.024 | 0.038 | 0.026 | 4.27 | 6.5E-05 | 3.93 | 2.5E-01 | 4.24 | 3.3E-05 | 0.95 | 0 | 3.36 | 1.8E-04 | 0.53 | 0 |
| STEAP4;ZNF804B | 7 | 88672600 | rs6465149 | T | C | 0.188 | 0.096 | 0.231 | 0.078 | 2.30 | 1.9E-04 | 3.41 | 5.6E-02 | 2.40 | 3.3E-05 | 0.56 | 0 | 1.71 | 6.5E-03 | 0.96 | 0 |
| KRAS;LMNTD1 | 12 | 25369798 | rs57251041 | A | G | 0.130 | 0.053 | 0.115 | 0.049 | 3.04 | 9.7E-05 | 2.82 | 1.6E-01 | 3.01 | 3.4E-05 | 0.92 | 0 | 2.62 | 6.3E-05 | 0.43 | 0 |
| DPPA4;LINC01205 | 3 | 109387060 | rs141872751 | G | A | 0.065 | 0.017 | 0.077 | 0.020 | 3.94 | 4.4E-04 | 11.05 | 1.7E-02 | 4.51 | 3.4E-05 | 0.34 | 0 | 3.87 | 5.8E-05 | 0.62 | 0 |
| NRG3;LINC02650 | 10 | 83599632 | rs74458842 | T | C | 0.091 | 0.033 | 0.077 | 0.017 | 3.39 | 1.8E-04 | 5.94 | 6.5E-02 | 3.59 | 3.5E-05 | 0.58 | 0 | 2.89 | 2.3E-04 | 0.78 | 0 |
| LINC02330;GALC | 14 | 87862194 | rs181231952 | A | G | 0.066 | 0.014 | 0.077 | 0.015 | 4.53 | 1.8E-04 | 6.83 | 7.2E-02 | 4.77 | 3.6E-05 | 0.72 | 0 | 3.31 | 1.1E-03 | 0.75 | 0 |
| CCSER1;LNCPRESS2 | 4 | 91805213 | rs142258961 | T | C | 0.039 | 0.006 | 0.038 | 0.006 | 7.70 | 1.2E-04 | 8.69 | 1.2E-01 | 7.82 | 3.6E-05 | 0.94 | 0 | 5.29 | 6.2E-04 | 0.95 | 0 |
| LINC02304;LINC02325 | 14 | 97418384 | rs78277874 | G | A | 0.032 | 0.004 | 0.038 | 0.006 | 11.14 | 6.0E-05 | 4.69 | 2.6E-01 | 9.70 | 3.7E-05 | 0.57 | 0 | 10.98 | 5.6E-06 | 0.74 | 0 |
| EFCAB5 | 17 | 30037016 | rs143673231 | T | A | 0.039 | 0.006 | 0.038 | 0.009 | 8.50 | 5.9E-05 | 4.52 | 3.0E-01 | 7.88 | 3.7E-05 | 0.68 | 0 | 5.40 | 5.6E-04 | 0.77 | 0 |
| SEZ6L | 22 | 26243032 | rs5761437 | G | C | 0.214 | 0.099 | 0.154 | 0.121 | 2.46 | 4.2E-05 | 1.63 | 4.8E-01 | 2.37 | 3.8E-05 | 0.57 | 0 | 1.81 | 2.1E-03 | 0.58 | 0 |
| CPT1A;MRPL21 | 11 | 68844064 | rs139866464 | A | C | 0.032 | 0.005 | 0.038 | 0.009 | 9.99 | 9.8E-05 | 6.17 | 1.7E-01 | 9.23 | 3.9E-05 | 0.74 | 0 | 5.93 | 7.4E-04 | 0.69 | 0 |
| LINC00533;LINC01623 | 6 | 28757075 | rs17398081 | C | G | 0.175 | 0.079 | 0.192 | 0.096 | 2.62 | 6.6E-05 | 1.91 | 2.7E-01 | 2.50 | 4.0E-05 | 0.62 | 0 | 1.97 | 9.3E-04 | 0.37 | 0 |
| ZPR1 | 11 | 116780747 | rs1942478 | G | T | 0.175 | 0.096 | 0.231 | 0.090 | 2.28 | 4.1E-04 | 4.37 | 2.2E-02 | 2.46 | 4.0E-05 | 0.34 | 0 | 2.16 | 9.3E-04 | 0.29 | 10 |
| UNC13A;MAP1S | 19 | 17700681 | rs28437574 | T | G | 0.208 | 0.346 | 0.192 | 0.370 | 0.46 | 2.3E-04 | 0.34 | 6.1E-02 | 0.44 | 4.0E-05 | 0.62 | 0 | 0.59 | 1.2E-03 | 0.44 | 0 |
| ERG | 21 | 38430644 | rs9974912 | T | C | 0.247 | 0.115 | 0.154 | 0.123 | 2.42 | 2.7E-05 | 1.26 | 7.1E-01 | 2.27 | 4.1E-05 | 0.32 | 0 | 1.64 | 2.7E-01 | 0.12 | 58 |
| DNAH8 | 6 | 38729478 | rs6458067 | T | C | 0.597 | 0.446 | 0.615 | 0.427 | 1.90 | 2.3E-04 | 2.33 | 6.6E-02 | 1.95 | 4.2E-05 | 0.68 | 0 | 1.63 | 4.9E-04 | 0.87 | 0 |
| SYCP1 | 1 | 114916759 | rs116660669 | A | G | 0.066 | 0.015 | 0.042 | 0.023 | 5.38 | 2.9E-05 | 1.67 | 6.5E-01 | 4.73 | 4.2E-05 | 0.34 | 0 | 3.12 | 4.4E-02 | 0.24 | 27 |
| ADGRV1 | 5 | 90751578 | rs138347163 | T | TAAC | 0.160 | 0.073 | 0.154 | 0.078 | 2.60 | 1.0E-04 | 2.30 | 1.9E-01 | 2.55 | 4.2E-05 | 0.86 | 0 | 2.28 | 7.5E-05 | 0.44 | 0 |
| KIAA1671 | 22 | 25039992 | rs11705626 | A | C | 0.214 | 0.115 | 0.231 | 0.108 | 2.27 | 2.8E-04 | 3.46 | 4.6E-02 | 2.38 | 4.2E-05 | 0.52 | 0 | 1.93 | 5.6E-04 | 0.61 | 0 |
| PACRG | 6 | 162950691 | rs187455695 | A | G | 0.039 | 0.006 | 0.038 | 0.006 | 8.13 | 1.4E-04 | 8.53 | 1.3E-01 | 8.18 | 4.2E-05 | 0.97 | 0 | 6.92 | 7.3E-05 | 0.77 | 0 |
| MYT1L;LINC01250 | 2 | 2694876 | rs58027731 | T | C | 0.120 | 0.049 | 0.115 | 0.048 | 3.05 | 9.6E-05 | 2.47 | 2.0E-01 | 2.96 | 4.3E-05 | 0.78 | 0 | 2.19 | 1.5E-03 | 0.58 | 0 |
| B3GAT2;OGFRL1 | 6 | 71245794 | rs116857090 | T | G | 0.052 | 0.010 | 0.077 | 0.015 | 5.46 | 2.1E-04 | 9.04 | 7.8E-02 | 5.79 | 4.3E-05 | 0.70 | 0 | 3.83 | 1.2E-03 | 0.85 | 0 |
| DLG2 | 11 | 85229144 | rs118120658 | T | G | 0.097 | 0.034 | 0.038 | 0.030 | 3.62 | 4.5E-05 | 1.95 | 5.7E-01 | 3.47 | 4.4E-05 | 0.61 | 0 | 3.02 | 7.7E-05 | 0.93 | 0 |
| MRPL21 | 11 | 68895031 | rs137898438 | A | G | 0.032 | 0.005 | 0.038 | 0.009 | 9.67 | 1.1E-04 | 6.17 | 1.7E-01 | 8.99 | 4.4E-05 | 0.76 | 0 | 5.75 | 8.4E-04 | 0.71 | 0 |
| IGHMBP2;MRGPRD | 11 | 68947262 | rs145432471 | C | T | 0.032 | 0.005 | 0.038 | 0.009 | 9.67 | 1.1E-04 | 6.17 | 1.7E-01 | 8.99 | 4.4E-05 | 0.76 | 0 | 5.75 | 8.4E-04 | 0.71 | 0 |
| PRRC2B | 9 | 131443142 | rs112421769 | A | T | 0.221 | 0.107 | 0.154 | 0.125 | 2.40 | 5.0E-05 | 1.63 | 4.6E-01 | 2.31 | 4.4E-05 | 0.58 | 0 | 1.79 | 2.0E-03 | 0.49 | 0 |
| KDM3B | 5 | 138365307 | rs55946397 | T | C | 0.059 | 0.016 | 0.077 | 0.015 | 3.97 | 6.6E-04 | 7.36 | 1.9E-02 | 4.44 | 4.5E-05 | 0.51 | 0 | 2.98 | 2.2E-03 | 0.58 | 0 |
| POMT1 | 9 | 131506797 | rs114317081 | A | G | 0.227 | 0.113 | 0.154 | 0.131 | 2.36 | 4.8E-05 | 1.55 | 5.2E-01 | 2.27 | 4.6E-05 | 0.55 | 0 | 1.75 | 2.4E-03 | 0.44 | 0 |
| B3GAT2 | 6 | 70932906 | rs148223971 | G | A | 0.032 | 0.004 | 0.038 | 0.015 | 12.25 | 3.9E-05 | 2.89 | 4.2E-01 | 9.51 | 4.6E-05 | 0.32 | 0 | 6.54 | 4.7E-04 | 0.37 | 0 |
| TTC29;MIR548G | 4 | 146971929 | rs78090635 | A | G | 0.078 | 0.022 | 0.038 | 0.018 | 4.47 | 3.9E-05 | 1.76 | 6.4E-01 | 4.14 | 4.6E-05 | 0.46 | 0 | 2.70 | 3.4E-03 | 0.34 | 0 |
| PABPC1P2;ACVR2A | 2 | 147367369 | rs80282306 | G | GA | 0.065 | 0.016 | 0.038 | 0.009 | 4.43 | 2.1E-04 | 10.94 | 7.3E-02 | 4.78 | 4.7E-05 | 0.52 | 0 | 3.24 | 1.7E-03 | 0.74 | 0 |
| CPA6 | 8 | 67648432 | rs422799 | C | T | 0.210 | 0.353 | 0.192 | 0.334 | 0.46 | 2.2E-04 | 0.33 | 7.5E-02 | 0.45 | 4.7E-05 | 0.60 | 0 | 0.48 | 1.5E-02 | 0.22 | 35 |
| PRDM5 | 4 | 120792381 | rs138061731 | G | A | 0.052 | 0.013 | 0.077 | 0.020 | 4.84 | 3.4E-04 | 6.36 | 5.2E-02 | 5.08 | 4.8E-05 | 0.79 | 0 | 4.55 | 4.1E-05 | 0.74 | 0 |
| PPP6R2 | 22 | 50419691 | rs8139476 | T | C | 0.467 | 0.324 | 0.423 | 0.334 | 1.96 | 2.1E-04 | 2.44 | 8.8E-02 | 2.00 | 4.8E-05 | 0.69 | 0 | 2.33 | 4.3E-02 | 0.08 | 68 |
| OBI1-AS1 | 13 | 78177406 | rs11619048 | T | C | 0.234 | 0.116 | 0.115 | 0.091 | 2.27 | 1.1E-04 | 2.61 | 2.0E-01 | 2.29 | 4.9E-05 | 0.86 | 0 | 2.10 | 4.9E-05 | 0.90 | 0 |
| TMEM248;SBDS | 7 | 66980308 | rs28585475 | A | G | 0.039 | 0.010 | 0.077 | 0.015 | 5.33 | 8.5E-04 | 11.15 | 1.6E-02 | 6.18 | 5.0E-05 | 0.51 | 0 | 3.90 | 1.7E-03 | 0.78 | 0 |

**Supplementary Table 2.** All variants associated with anthracycline-related arrhythmia and cardiac events at the chr19 *CHST8*:*KCTD15 and* chr8 *MMP16* loci, p<5E-06 in meta-analyses. CHR, chromosome; BP, Base-pair coordinate Hg38, A1, Allele 1; A2, Allele 2; OR, Odds Ratio; Q, p-value for Cochran’s Q statistic; I, I^2^ heterogeneity index (0-100 scale). Allele freq, Allele frequency in cases with Arrhythmia (ARR) and without Arrhythmia (no ARR) in MER and Tapestry datasets.

| r | BP | rsID | A1 | A2 | Allele frequency arrhythmia |  |  |  | MER arrhythmia |  | Tapestry arrhythmia |  | Meta-analysis arrhythmia |  |  |  | Meta-analysis cardiac event |  |  |
| --- | --- | --- | --- | --- | --- | --- | --- | --- | --- | --- | --- | --- | --- | --- | --- | --- | --- | --- | --- |
|  |  |  |  |  | MER ARR | MER No ARR | Tapestry ARR | Tapestry No ARR | P | OR | P | OR | P | OR | Q | I | P | OR | Q |
| 8 | 88457185 | rs1094199 | T | C | 0.079 | 0.019 | 0.038 | 0.015 | 1.09E-06 | 6.08 | 0.150 | 5.90 | 3.72E-07 | 6.07 | 0.98 | 0 | 1.98E-08 | 6.15 | 0.52 |
| 8 | 88462911 | rs117646114 | A | G | 0.071 | 0.016 | 0.038 | 0.017 | 1.24E-06 | 6.47 | 0.168 | 5.38 | 4.68E-07 | 6.37 | 0.89 | 0 | 1.34E-06 | 5.25 | 0.49 |
| 8 | 88471665 | rs1685555 | G | A | 0.086 | 0.023 | 0.038 | 0.020 | 2.78E-06 | 5.24 | 0.236 | 4.13 | 1.36E-06 | 5.14 | 0.85 | 0 | 1.97E-07 | 4.95 | 0.38 |
| 8 | 88430321 | rs61433177 | C | A | 0.084 | 0.023 | 0.038 | 0.020 | 3.33E-06 | 5.16 | 0.236 | 4.13 | 1.62E-06 | 5.07 | 0.86 | 0 | 2.29E-07 | 4.90 | 0.39 |
| 8 | 88360812 | rs60298754 | T | C | 0.078 | 0.020 | 0.038 | 0.017 | 4.19E-06 | 5.39 | 0.209 | 4.56 | 1.85E-06 | 5.31 | 0.89 | 0 | 1.05E-06 | 4.81 | 0.45 |
| 8 | 88368978 | rs73277117 | C | A | 0.078 | 0.020 | 0.038 | 0.017 | 4.19E-06 | 5.39 | 0.209 | 4.56 | 1.85E-06 | 5.31 | 0.89 | 0 | 1.05E-06 | 4.81 | 0.45 |
| 8 | 88383579 | rs73277199 | G | A | 0.078 | 0.020 | 0.038 | 0.017 | 4.19E-06 | 5.39 | 0.209 | 4.56 | 1.85E-06 | 5.31 | 0.89 | 0 | 1.05E-06 | 4.81 | 0.45 |
| 8 | 88635459 | rs80023829 | A | T | 0.065 | 0.016 | 0.038 | 0.018 | 5.88E-06 | 5.98 | 0.169 | 5.35 | 2.20E-06 | 5.91 | 0.93 | 0 | 3.72E-06 | 5.04 | 0.63 |
| 8 | 88676432 | rs75966776 | G | A | 0.065 | 0.016 | 0.038 | 0.018 | 5.88E-06 | 5.98 | 0.169 | 5.35 | 2.20E-06 | 5.91 | 0.93 | 0 | 3.72E-06 | 5.04 | 0.63 |
| 8 | 88331620 | rs80039465 | G | A | 0.065 | 0.016 | 0.038 | 0.012 | 9.02E-06 | 5.80 | 0.106 | 7.77 | 2.36E-06 | 5.96 | 0.83 | 0 | 4.34E-06 | 5.03 | 0.81 |
| 8 | 88364075 | rs17730333 | G | A | 0.065 | 0.016 | 0.038 | 0.015 | 9.02E-06 | 5.80 | 0.142 | 6.12 | 2.94E-06 | 5.83 | 0.97 | 0 | 5.95E-06 | 4.89 | 0.62 |
| 8 | 88426720 | rs79676922 | G | A | 0.078 | 0.022 | 0.038 | 0.017 | 8.79E-06 | 5.03 | 0.168 | 5.38 | 3.26E-06 | 5.06 | 0.96 | 0 | 3.13E-07 | 5.01 | 0.52 |
| 8 | 88369785 | rs117758893 | G | A | 0.071 | 0.019 | 0.038 | 0.015 | 1.11E-05 | 5.27 | 0.142 | 6.12 | 3.61E-06 | 5.34 | 0.91 | 0 | 1.42E-06 | 4.94 | 0.62 |
| 8 | 88376375 | rs76034445 | C | G | 0.071 | 0.019 | 0.038 | 0.015 | 1.11E-05 | 5.27 | 0.142 | 6.12 | 3.61E-06 | 5.34 | 0.91 | 0 | 1.42E-06 | 4.94 | 0.62 |
| 8 | 88404021 | rs55807042 | T | C | 0.078 | 0.022 | 0.038 | 0.020 | 1.14E-05 | 4.90 | 0.236 | 4.13 | 5.51E-06 | 4.83 | 0.89 | 0 | 6.20E-07 | 4.76 | 0.40 |
| 8 | 88429426 | rs77948155 | T | C | 0.078 | 0.022 | 0.038 | 0.020 | 1.14E-05 | 4.90 | 0.236 | 4.13 | 5.51E-06 | 4.83 | 0.89 | 0 | 6.20E-07 | 4.76 | 0.40 |
| 8 | 88449678 | rs60484072 | T | A | 0.078 | 0.022 | 0.038 | 0.020 | 1.14E-05 | 4.90 | 0.236 | 4.13 | 5.51E-06 | 4.83 | 0.89 | 0 | 6.20E-07 | 4.76 | 0.40 |
| 8 | 88331580 | rs73692229 | C | T | 0.078 | 0.023 | 0.038 | 0.015 | 2.33E-05 | 4.59 | 0.168 | 5.41 | 8.66E-06 | 4.65 | 0.90 | 0 | 9.40E-07 | 4.61 | 0.62 |
| 8 | 88328028 | rs28907872 | A | C | 0.078 | 0.023 | 0.038 | 0.015 | 2.34E-05 | 4.61 | 0.168 | 5.41 | 8.71E-06 | 4.67 | 0.90 | 0 | 8.84E-07 | 4.66 | 0.61 |
| 8 | 88328072 | rs28907871 | T | C | 0.078 | 0.023 | 0.038 | 0.015 | 2.34E-05 | 4.61 | 0.168 | 5.41 | 8.71E-06 | 4.67 | 0.90 | 0 | 8.84E-07 | 4.66 | 0.61 |
| 8 | 88325008 | rs73692228 | T | A | 0.071 | 0.020 | 0.038 | 0.012 | 3.71E-05 | 4.84 | 0.106 | 7.77 | 1.01E-05 | 5.03 | 0.72 | 0 | 5.52E-07 | 5.22 | 0.78 |
| 8 | 88403848 | rs80287386 | A | G | 0.071 | 0.021 | 0.038 | 0.017 | 2.99E-05 | 4.77 | 0.168 | 5.38 | 1.11E-05 | 4.82 | 0.92 | 0 | 8.58E-07 | 4.87 | 0.54 |
| 8 | 88403967 | rs78204053 | G | A | 0.071 | 0.021 | 0.038 | 0.017 | 2.99E-05 | 4.77 | 0.168 | 5.38 | 1.11E-05 | 4.82 | 0.92 | 0 | 8.58E-07 | 4.87 | 0.54 |
| 8 | 88328299 | rs3840718 | CAA | C | 0.072 | 0.023 | 0.038 | 0.015 | 8.73E-05 | 4.31 | 0.168 | 5.41 | 3.25E-05 | 4.39 | 0.86 | 0 | 2.73E-06 | 4.47 | 0.63 |
| 9 | 33790783 | rs12151014 | A | G | 0.195 | 0.069 | 0.192 | 0.079 | 1.17E-07 | 3.52 | 0.024 | 4.27 | 9.01E-09 | 3.61 | 0.78 | 0 | 2.73E-07 | 2.84 | 0.63 |
| 9 | 33792898 | rs77793167 | A | G | 0.195 | 0.069 | 0.192 | 0.079 | 1.22E-07 | 3.52 | 0.024 | 4.27 | 9.38E-09 | 3.60 | 0.78 | 0 | 2.85E-07 | 2.84 | 0.64 |
| 9 | 33796072 | rs62101381 | A | C | 0.195 | 0.069 | 0.192 | 0.079 | 1.22E-07 | 3.52 | 0.025 | 4.24 | 9.55E-09 | 3.60 | 0.78 | 0 | 2.95E-07 | 2.83 | 0.62 |
| 9 | 33796347 | rs141116709 | T | C | 0.195 | 0.069 | 0.192 | 0.079 | 1.27E-07 | 3.51 | 0.024 | 4.24 | 9.47E-09 | 3.59 | 0.78 | 0 | 3.06E-07 | 2.83 | 0.62 |
| 9 | 33802527 | rs62101382 | C | A | 0.195 | 0.070 | 0.192 | 0.079 | 1.49E-07 | 3.48 | 0.025 | 4.24 | 1.18E-08 | 3.57 | 0.77 | 0 | 3.62E-07 | 2.81 | 0.63 |
| 9 | 33803888 | rs80246986 | G | A | 0.195 | 0.070 | 0.154 | 0.079 | 1.49E-07 | 3.48 | 0.095 | 3.04 | 3.64E-08 | 3.43 | 0.85 | 0 | 9.35E-07 | 2.73 | 0.36 |
| 9 | 33791805 | rs8108556 | C | T | 0.195 | 0.070 | 0.192 | 0.079 | 2.26E-07 | 3.43 | 0.024 | 4.27 | 1.76E-08 | 3.53 | 0.75 | 0 | 4.21E-07 | 2.80 | 0.65 |
| 9 | 33792516 | rs117523590 | C | T | 0.195 | 0.070 | 0.192 | 0.079 | 2.26E-07 | 3.43 | 0.024 | 4.27 | 1.76E-08 | 3.53 | 0.75 | 0 | 4.21E-07 | 2.80 | 0.65 |
| 9 | 33789921 | rs10412871 | A | G | 0.195 | 0.071 | 0.192 | 0.082 | 2.90E-07 | 3.39 | 0.025 | 4.26 | 2.26E-08 | 3.49 | 0.74 | 0 | 5.70E-07 | 2.76 | 0.67 |
| 9 | 33789264 | rs13345814 | G | A | 0.195 | 0.071 | 0.192 | 0.082 | 3.38E-07 | 3.37 | 0.025 | 4.26 | 2.63E-08 | 3.47 | 0.73 | 0 | 6.74E-07 | 2.75 | 0.68 |
| 9 | 33789255 | rs61525454 | G | GTC | 0.195 | 0.072 | 0.192 | 0.082 | 4.08E-07 | 3.34 | 0.025 | 4.26 | 3.18E-08 | 3.44 | 0.72 | 0 | 8.21E-07 | 2.72 | 0.69 |
| 9 | 33789276 | rs10406150 | A | G | 0.195 | 0.072 | 0.192 | 0.082 | 4.08E-07 | 3.34 | 0.025 | 4.26 | 3.18E-08 | 3.44 | 0.72 | 0 | 8.21E-07 | 2.72 | 0.69 |

**Supplementary Figure 1:**
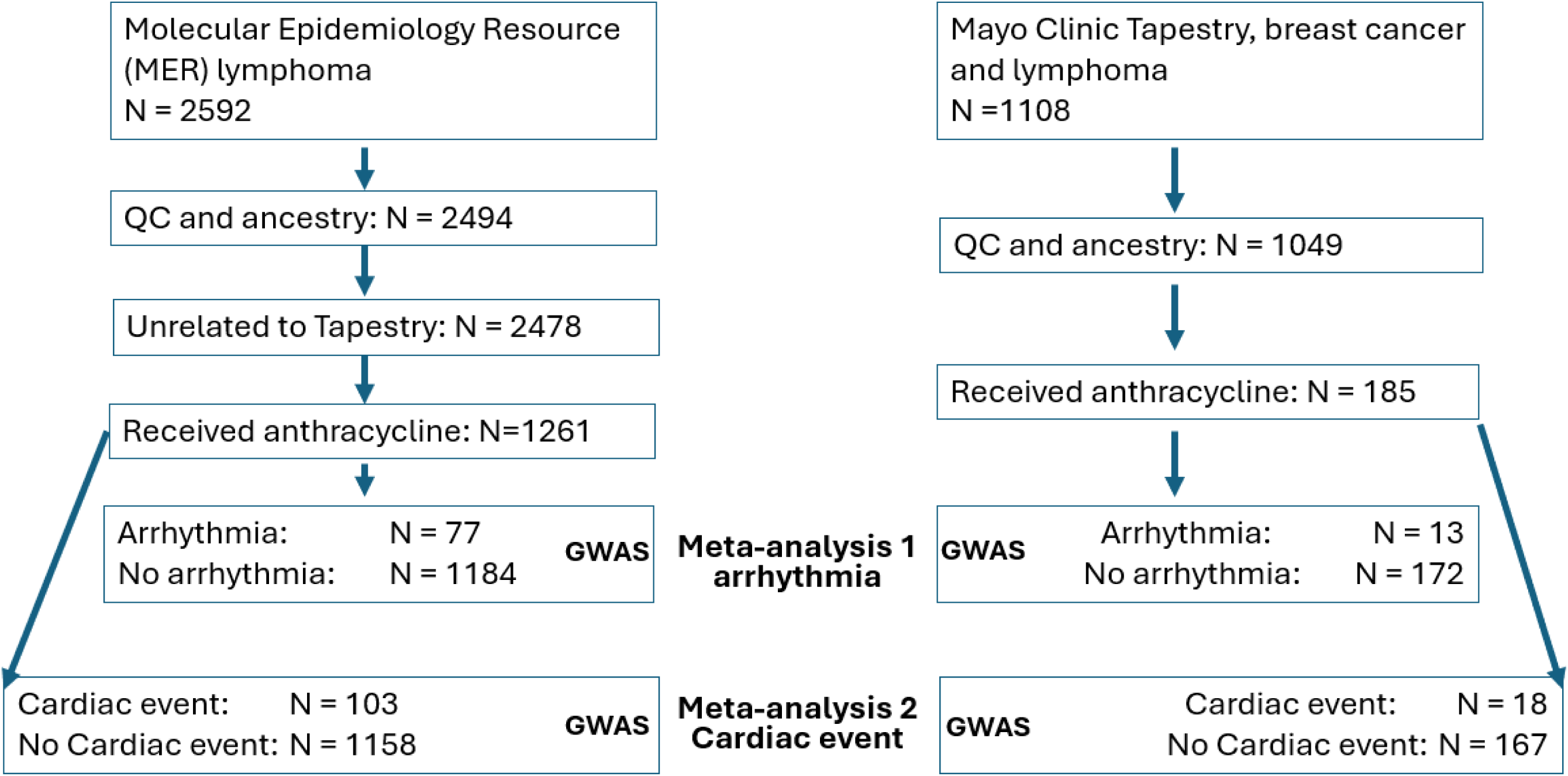
Consort diagram of genotyping QC and numbers of each sample.

**Supplementary Figure 2:**
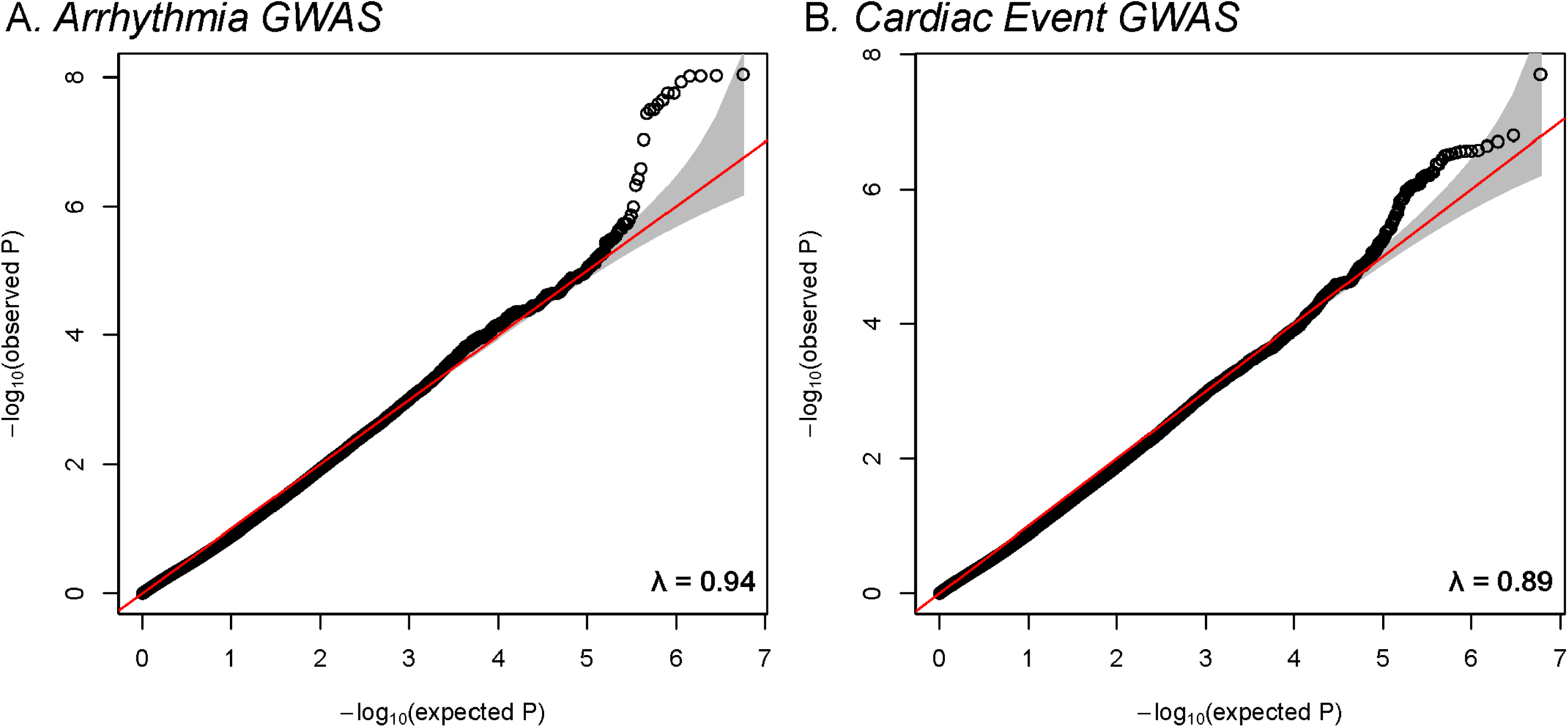
Meta-analysis QQ plots of arrhythmia and cardiac event outcomes. (A) Meta-analysis of MER and Tapestry GWAS datasets with outcome of arrhythmia, lambda = 0.94. (B) Meta-analysis of MER and Tapestry GWAS datasets with outcome of cardiac event, lambda = 0.89.

**Supplementary Figure 3:**
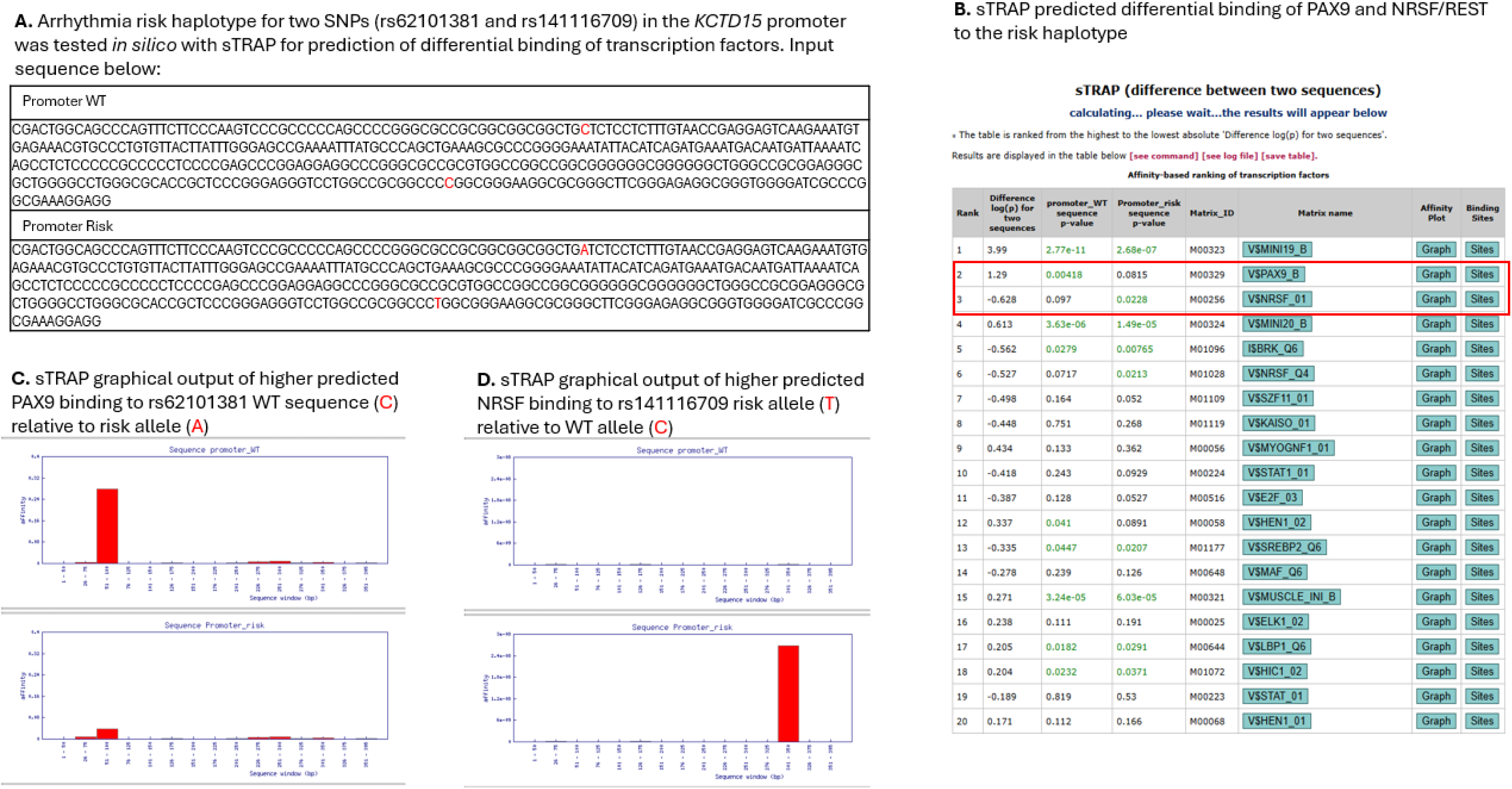
Predicted differential binding of NRSF/REST transcription factor to the associated variants in the *KCTD15* promoter.

## Project Generation

### Mayo Clinic

PG Leadership Team: James Cerhan, Fergus Couch, Janet Olson

Statistical Genetics and Bioinformatics: Nicholas Larson, Zachary S. Fredericksen Laboratory Operations: Mine Cicek

### Regeneron Genetics Center (Tarrytown, NY, USA)

RGC Management & Leadership Team: Aris Baras, Gonçalo Abecasis, Adolfo Ferrando, Giovanni Coppola, Andrew Deubler, Luca A Lotta, John D Overton, Jeffrey G Reid, Alan Shuldiner, Katherine Siminovitch, Jason Portnoy, Marcus B Jones, Lyndon Mitnaul, Alison Fenney, Jonathan Marchini, Manuel Allen Revez Ferreira, Maya Ghoussaini, Mona Nafde, William Salerno, Cristen Willer, Lourdes Crane.

Sequencing & Lab Operations: John D Overton, Christina Beechert, Erin Fuller, Laura M Cremona, Eugene Kalyuskin, Hang Du, Caitlin Forsythe, Zhenhua Gu, Kristy Guevara, Michael Lattari, Alexander Lopez, Kia Manoochehri, Prathyusha Challa, Manasi Pradhan, Raymond Reynoso, Ricardo Schiavo, Maria Sotiropoulos Padilla, Chenggu Wang, Sarah E Wolf, Hang Du, Kristy Guevara.

Genome Informatics & Data Engineering: Jeffrey G Reid, Mona Nafde, Manan Goyal, George Mitra, Sanjay Sreeram, Rouel Lanche, Vrushali Mahajan, Sai Lakshmi Vasireddy, Gisu Eom, Krishna Pawan Punuru, Sujit Gokhale, Benjamin Sultan, Pooja Mule, Mudasar Sarwar, Muhammad Aqeel, Xiaodong Bai, Lance Zhang, Sean O’Keeffe, Razvan Panea, Evan Edelstein, Ayesha Rasool, William Salerno, Evan K Maxwell, Boris Boutkov, Alexander Gorovits, Ju Guan, Lukas Habegger, Alicia Hawes, Olga Krasheninina, Samantha Zarate, Adam J Mansfield, Lukas Habegger.

Analytical Genetics and Data Science: Gonçalo Abecasis, Manuel Allen Revez Ferreira, Joshua Backman, Kathy Burch, Adrian Campos, Liron Ganel, Sheila Gaynor, Benjamin Geraghty, Arkopravo Ghosh, Salvador Romero Martinez, Christopher Gillies, Lauren Gurski, Eric Jorgenson, Tyler Joseph, Michael Kessler, Jack Kosmicki, Adam Locke, Priyanka Nakka, Jonathan Marchini, Karl Landheer, Olivier Delaneau, Maya Ghoussaini, Anthony Marcketta, Joelle Mbatchou, Arden Moscati, Anita Pandit, Jonathan Ross, Carlo Sidore, Eli Stahl, Timothy Thornton, Sailaja Vedantam, Rujin Wang, Kuan-Han Wu, Bin Ye, Blair Zhang, Andrey Ziyatdinov, Yuxin Zou, Jingning Zhang, Kyoko Watanabe, Mira Tang, Frank Wendt, Suganthi Balasubramanian, Suying Bao, Kathie Sun, Chuanyi Zhang, Sean Yu, Aaron Zhang, David Corrigan, Dhruv Shidhaye, Chen Wang, Keyrun Adhikari, Alexander Lachmann.

Therapeutic Area Genetics: Adolfo Ferrando, Giovanni Coppola, Luca A. Lotta, Alan Shuldiner, Katherine Siminovitch, Brian Hobbs, Jon Silver, William Palmer, Rita Guerreiro, Amit Joshi, Antoine Baldassari, Cristen Willer, Sarah Graham, Ernst Mayerhofer, Erola Pairo Castineira, Mary Haas, Niek Verweij, George Hindy, Jonas Bovijn, Tanima De, Luanluan Sun, Olukayode Sosina, Arthur Gilly, Peter Dornbos, Juan Rodriguez-Flores, Moeen Riaz, Manav Kapoor, Gannie Tzoneva, Momodou W Jallow, Anna Alkelai, Ariane Ayer, Veera Rajagopal, Sahar Gelfman, Vijay Kumar, Jacqueline Otto, Jose Bras, Silvia Alvarez, Jessie Brown, Hossein Khiabanian, Joana Revez, Kimberly Skead, Valentina Zavala, Jae Soon Sul, Lei Chen, Sam Choi, Amy Damask, Nan Lin, Charles Paulding, Sameer Malhotra, Joseph Herman.

Research Program Management & Strategic Initiatives: Marcus B Jones, Michelle G LeBlanc, Nadia Rana, Jennifer Rico-Varela, Jaimee Hernandez, Larizbeth Romero, Ashley Paynter.

Senior Partnerships & Business Operations: Randi Schwartz, Lourdes Crane, Alison Fenney, Jody Hankins, Anna Han, Samuel Hart, Ryan Smith.

Business Operations & Administrative Coordinators: Ann Perez-Beals, Gina Solari, Johannie Rivera-Picart, Michelle Pagan, Sunilbe Siceron.

### Sources of funding

Research reported in this publication was supported by the National Heart Lung and Blood Institute of the National Institutes of Health under Award Number R01 HL169268 and by the Department of Defense under award number W81XWH-22-1-0288/0289 (PR210385). MER supported by the National Cancer Institute of the National Institutes of Health under Awards P50 CA97274 and U01 CA195568. Project Generation was supported in part by the Center for Individualized Medicine, Mayo Clinic.

Genomic data were provided by the Tapestry study supported by the Mayo Clinic Center for Individualized Medicine. We thank all of the participants of the Tapestry study.

## Disclosures

### Data availability

Project Generation data were used under license, thus are not publicly available due to use restrictions. Reasonable requests for Project Generation data from qualified researchers may be directed to, Dr. Jim Cerhan. Access to de-identified data will require a legal agreement and the permission of Regeneron.

Tapestry data were used under license, thus are not publicly available due to use restrictions. Reasonable requests for Tapestry data from qualified researchers may be directed to, Dr. Konstantinos Lazaridis. Access to de-identified data will require a legal agreement and the permission of Helix, Inc.

### Appendix: Tapestry Cohort Author List

Ahmad H. Ali, M.B.B.S. 1; Sebastian M. Armasu, M.S.; M.A. 2; Arjun P. Athreya, Ph.D.; M.S. 3; Sanjay P. Bagaria, M.D. 4; Lorelei A. Bandel, M.B.A. 5; Jamie K. Bruflat, M.S. 5; Michelle L. Bublitz, M.H.A., CCRP 5; Brittany N. Burnap 5; Elliot J. Cahn 2; Wenan Chen, Ph.D. 2; Blake J. Clermont 5; Jan B. Egan, Ph.D. 6; Idara U. Ekpoh, M.B.A. 6; Nate P. Erwin M.B.A., CSM 7; Jacob A. Frank 2; Courtney B. Graddy, M.H.A. 8; Leila A. Jones, Ph.D. 5; Suneetha Kaggal 2; Mary E. Karow, M.S. 5, 9; Jennifer L. Kemppainen, M.S., CGC 5; Eric W. Klee, Ph.D. 2,5,10; Nicholas B. Larson, Ph.D., M.S. 2; Konstantinos N. Lazaridis, M.D. 1,5; Jon W. Leitzke 2; Tony C. Luehrs, M.S. 2; Tammy M. McAllister, M.A. 5; Joel Morales-Rosado, M.D. 5,11; Olson, Rory, Ph.D. 5; Victor E. Ortega, M.D., Ph.D. 6; Melanie M. Peterson, M.S. 5; Robert J. Pignolo, M.D., Ph.D. 12,13,14; Filippo Pinto e Vairo, M.D., Ph.D. 5; John J. Presutti, D.O., FAAFP 15; Stephanie Safgren, Ph.D. 16; Jewel J. Samadder, M.D. 1; Aleksandar Sekulic, M.D., Ph.D. 17; Hugues Sicotte, Ph.D. 2; Tan-Arroyo, Jennifer, Ph.D. 5; Shulan Tian, Ph.D., M.S. 2; Kristine M. Tree, CSM 18; Robert A. Vierkant, M.S. 2; T’Nita S. Walker 5; Tammy A. Wilson 5

1. Division of Gastroenterology and Hepatology, College of Medicine, Mayo Clinic, Rochester, MN, USA
2. Department of Quantitative Health Sciences, Mayo Clinic, Rochester, MN, USA
3. Department of Psychiatry and Psychology, Mayo Clinic, Rochester, MN, USA
4. Division of Surgical Oncology, Mayo Clinic, Jacksonville, FL, USA
5. Center for Individualized Medicine, Mayo Clinic, Rochester, MN, USA
6. Center for Individualized Medicine, Mayo Clinic, Scottsdale, AZ, USA
7. Research Strategic Transformation, Mayo Clinic, Rochester, MN, USA
8. Center for Individualized Medicine, Mayo Clinic, Jacksonville, FL, USA
9. Mayo Clinic GeneGuide Laboratory, Mayo Clinic, Rochester, MN, USA
10. Department of Clinical Genomics, Mayo Clinic, Rochester, MN, USA
11. Now at: Vanderbilt University School of Medicine, 861 18st Ave S # D3300, Nashville, TN
12. Department of Medicine, Mayo Clinic, Rochester, MN, USA
13. Robert and Arlene Kogod Center on Aging, Mayo Clinic, Rochester, MN, USA
14. Department of Physiology and Biomedical Engineering, Mayo Clinic, Rochester, MN, USA
15. Department of Family Medicine, Mayo Clinic, Jacksonville, FL, USA
16. Department of Oncology, Division of Oncology Research, Mayo Clinic, Rochester, MN, USA
17. Department of Dermatology, Mayo Clinic, Scottsdale, AZ, USA
18. Research Strategic Transformation, Mayo Clinic, Rochester, MN, USA

